# A rapid review of what innovative workforce models have helped to rapidly grow capacity for community care to help older adults leave hospital

**DOI:** 10.1101/2022.08.05.22278310

**Authors:** Helen Morgan, Alison Weightman, Kate J Lifford, Lydia Searchfield, Mala Mann, Freya Davies, Ruth Lewis, Alison Cooper, Adrian Edwards

**Affiliations:** Specialist Unit for Review Evidence, Cardiff University, Wales, United Kingdom; Wales COVID-19 Evidence Centre, Wales, United Kingdom

## Abstract

Older adults who have undergone treatment as hospital inpatients and are now medically fit for discharge back into the community may require additional care to support that transition. Prolonged hospital admissions can also have risks including functional decline, dependency and the risk of hospital acquired infections.

The aim of this rapid review was to review the research evidence for the effectiveness of workforce models in the community that may be able to rapidly grow capacity for community care and help older adults leave hospital.

19 studies were included: 11 systematic reviews and 8 UK primary studies not included in the reviews (4 quantitative study designs, 1 case study,1 mixed method study and 2 qualitative studies). The 19 studies evaluated 5 different intervention areas and a range of outcomes including: hospital length of stay; bed day rates, days to early supported discharge, delayed transfers of care (DTOCs); episode length of care; mortality; readmission; and carer, patient or staff perceptions.

Intervention areas that were studied the most were: Early Supported Discharge and Transitional care/Continuity of Care. Nine recent UK studies from the systematic reviews describing these interventions were analysed separately for data on outcomes and workforce components.

It is difficult to draw firm conclusions due to the limited evidence from a UK setting, and low quality of included studies. There is insufficient information to propose an optimum service design, but the evidence does suggest that interventions that are more comprehensive (covering a range of different components) and more intensive are more likely to be effective.

Further research is needed to evaluate the effectiveness of workforce models introduced to rapidly grow capacity for community care to help older adults leave hospital in the UK setting. The Early Supported Discharge and Transitional Care models hold some promise.

Wales COVID-19 Evidence Centre (WCEC) Rapid Review

Report number — RR00039 (July 2022)

Rapid Review Details
**Review conducted by:**
Specialist Unit for Review Evidence (SURE), Cardiff University
**Review Team:**
Helen Morgan, Alison Weightman, Kate Lifford, Lydia Searchfield, Mala Mann, Freya Davies
**Review submitted to the WCEC on:**
15^th^ June 2022
**Stakeholder consultation meeting:**
21^st^ June 2022
**Rapid Review report issued by the WCEC on:**
July 2022
**WCEC Team:**
Alison Cooper, Ruth Lewis, Adrian Edwards, Micaela Gal, Jane Greenwell
**This review should be cited as:**
RR00039. Wales COVID-19 Evidence Centre. A rapid review of what innovative workforce models have helped to rapidly grow capacity for community care to help older adults leave hospital. July 2022

Disclaimer The views expressed in this publication are those of the authors, not necessarily Health and Care Research Wales. The WCEC and authors of this work declare that they have no conflict of interest.

TOPLINE SUMMARY

What is a Rapid Review?
Our rapid reviews use a variation of the systematic review approach, abbreviating or omitting some components to generate the evidence to inform stakeholders promptly whilst maintaining attention to bias. They follow the methodological recommendations and minimum standards for conducting and reporting rapid reviews, including a structured protocol, systematic search, screening, data extraction, quality appraisal, and evidence synthesis to answer a specific question and identify key research gaps. This Rapid Review was undertaken in **four weeks** to inform an urgent policy priority. Existing systematic reviews and UK primary studies were prioritised.

Who is this summary for?
The review was requested urgently from Social Care Wales and Health Education and Improvement Wales to help inform the potential workforce planning and service responses across social care and health services to create additional community care capacity to support individuals to live at home, and support timely discharge from hospital.

Background / Aim of Rapid Review
Older adults who have undergone treatment as hospital inpatients and are now **medically fit** for discharge back into the community may require **additional care to support that transition**. Prolonged hospital admissions can also have risks including functional decline, dependency and the risk of hospital acquired infections. The aim of this review was to review the research evidence for the effectiveness of **workforce models in the community** that may be able to rapidly grow capacity for community care and help older adults leave hospital.

Key Findings
**19 studies** were included: 11 systematic reviews and 8 UK primary studies not included in the reviews (4 quantitative study designs, 1 case study,1 mixed method study and 2 qualitative studies).

Extent of the evidence base

- The 19 studies evaluated **5 different intervention areas** and a **range of outcomes** including: hospital length of stay; bed day rates, days to early supported discharge, delayed transfers of care (DTOCs); episode length of care; mortality; readmission; and carer, patient or staff perceptions.
- Intervention areas that were studied the most were: **Early Supported Discharge** and **Transitional care/Continuity of Care**. Nine recent UK studies from the systematic reviews describing these interventions were analysed separately for data on outcomes and workforce components.

Recency of the evidence base

- Articles were published **2016-2022**
- The UK studies extracted from the systematic reviews were published from 2000-2017.

Evidence of effectiveness

- The body of systematic review evidence for **Early Supported Discharge** is consistent and indicates that early supported discharge **reduces length of hospital stay** although the recent UK studies did not show a consistent pattern.
- The body of systematic review evidence for **Transitional care/Continuity of care** is fairly consistent and tended to demonstrate **a reduction in readmissions** although the recent UK studies did not show a consistent pattern.
- All the **interventions** evaluated were **multi-disciplinary**.
- **Geographical location** adds to the **heterogeneity** in intervention designs and outcomes.
- **Staff perceptions,** reported by Baxter et al. (2020), informing how safe transitions can be facilitated include: **getting to know the patient**; building **relationships** within and across **teams** and **bridging gaps** within the **system.**

Policy Implications

- It is difficult to draw firm conclusions due to the limited evidence from a UK setting, and low quality of included studies.
- There is insufficient information to propose an optimum service design but the evidence does suggest that **interventions that are more comprehensive** (covering a range of different components) and **more intensive** are more likely to be effective.
- **Further research is needed** to evaluate the effectiveness of workforce models introduced to rapidly grow capacity for community care to help older adults leave hospital in the UK setting.
- The Early Supported Discharge and Transitional Care models hold some promise.

Strength of Evidence

- None of the systematic reviews was rated as high quality.
- Of the 8 primary studies identified that were not included in the systematic reviews, 1 primary quantitative study and one primary qualitative study were rated as high quality.
- The 9 UK studies identified within the systematic reviews on Early Supported Discharge and Transitional Care/Continuity of Care were not quality appraised.

## 1. BACKGROUND

This Rapid Review is being conducted as part of the Wales COVID-19 Evidence Centre Work Programme. The review was requested urgently from Social Care Wales and Health Education and Improvement Wales to help inform the potential workforce planning and service responses across social care and health services to create additional community care capacity to support individuals to live at home, and support timely discharge from hospital.

### 1.1 Purpose of this review

Older adults that have undergone treatment as a hospital inpatient and are now medically fit for discharge back into the community may require additional care to support that transition. Prolonged hospital admissions can also have risks including functional decline, dependency and the risk of hospital <u>acquired infections.</u> The aim of this review was to understand the evidence for workforce models in the community to rapidly grow capacity for community care to help older adults leave hospital.

## 2. RESULTS

### 2.1 Overview of the Evidence Base

From screening 199 records, 19 studies met the inclusion criteria and were included in this review. Those 19 studies included: 11 systematic reviews, 4 quantitative study designs, 1 case study,1 mixed method study and 2 qualitative studies. None of the separately identified primary studies were included in any of the systematic reviews.

The 11 systematic reviews included internationally conducted primary studies and all except for one review (Mabire et al. 2016) included at least one study conducted in the UK. Of the 7 included primary studies, 6 were conducted in England and one in Scotland. The case study was conducted in the Hywel Dda University Health Board region.

Four of the systematic reviews were Cochrane reviews (Gonçalves-Bradley et al. 2017, 2022; Handoll et al. 2021 Langhorne et al. 2017), the other 7 systematic reviews and 5 of the primary studies were published as journal articles. Home Based Bridging Care Project Evaluation (2022), Jones et al. (2022) and National Team for Inclusion (2019) were ‘grey literature’ i.e., reports published outside of traditional commercial publishing^1^.

The 19 studies evaluated five different intervention areas (Table 1) and a range of outcomes including: hospital length of stay; bed day rates, days to early supported discharge, delayed transfers of care (DTOCs); episode length of care; mortality; readmission; as well as carer, patient or staff perceptions. The intervention areas that were studied the most were Transitional care/Continuity of care (Baxter et al. 2020; Facchinetti et al. 2020; Lee et al. 2022; Meulenbroeks et al. 2021; Rasmussen et al. 2021; Tomlinson et al. 2020) and Early Supported Discharge^2^ (Davies et al. 2019, Fisher et al. 2020; Gonçalves-Bradley et al. 2017; Home Based Bridging Care Project Evaluation 2022, Langhorne et al. 2017; Williams et al. 2022). Where outcomes for each of these intervention areas were reported by 3 or more studies then a summary of those outcomes are provided at the end of the relevant sections. Further details are also provided in tables 1-3 with table 1 displaying the evidence base by intervention and outcomes.

**Table 1.**
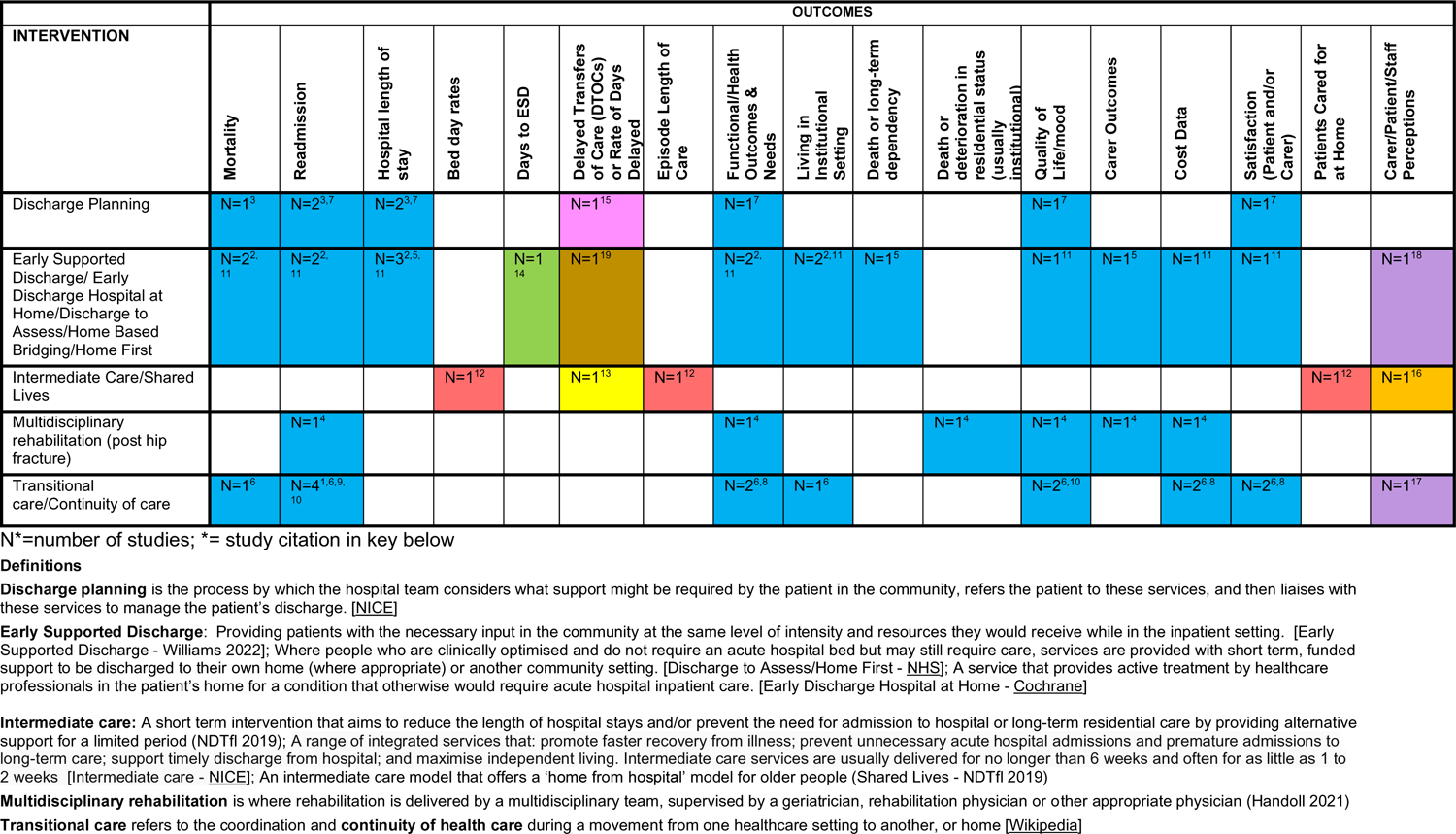
Overview of Evidence Base by Intervention and Outcomes

The quality of the systematic reviews was assessed using the <u>AMSTAR 2</u> quality appraisal tool. According to AMSTAR 2, quality of 3 of the systematic reviews (Gonçalves-Bradley et al. 2022; Handoll et al. 2021; Tomlinson et al. 2020) were rated as moderate; one was rated as low (Rasmussen et al. 2021) and 7 as critically low (Facchinetti et al. 2020; Gonçalves-Bradley et al. 2017; Langhorne et al. 2017; Lee et al. 2022; Mabire et al. 2016; Meulenbroeks et al. 2021 Williams et al. 2022). The primary studies were appraised using the appropriate quality appraisal form (see Section 5.8). The quality of 3 of the primary quantitative studies (Elston et al. 2022; Jones et al., 2022; Levin and Crighton 2019) was rated as moderate; Fisher et al. (2020) was rated as high. The mixed methods study was rated as low quality, principally as this was a pilot study. The qualitative studies, Baxter et al. (2020) and Davis et al. (2019) were rated as high and moderate quality respectively. The case study (Home Based Bridging Care, 2022) was not appraised as no appropriate quality appraisal tool exists to appraise this level of evidence when the population comprises a region. Further details are provided in the quality appraisal tables (Section 6.3).

#### Key

| Study Type | Citation | Quality Rating |
| --- | --- | --- |
| Systematic Review | 1. Facchinetti 2020 | Critically low |
|  | 2. Gonçalves-Bradley 2017 | Critically low |
|  | 3. Gonçalves-Bradley 2022 | Moderate |
|  | 4. Handoll 2021 | Moderate |
|  | 5. Langhorne 2017 | Critically low |
|  | 6. Lee 2022 | Critically low |
|  | 7. Mabire 2016 | Critically low |
|  | 8. Meulenbroeks 2021 | Critically low |
|  | 9. Rasmussen 2021 | Low |
|  | 10. Tomlinson 2020 | Low |
|  | 11. Williams 2022 | Low |
| Controlled Before & After | 12. Elston 2022 | Moderate |
| Interrupted Time Series | 13. Levin 2019 | Moderate |
| Cohort | 14. Fisher 2020 | High |
| Cross-sectional | 15. Jones 2022 | Moderate |
| Mixed methods | 16. National Development Team for Inclusion (NDTfi) 2019 | Low |
| Qualitative study | 17. Baxter 2020 | High |
|  | 18. Davis 2019 | Moderate |
| Case study | 19. Home Based Bridging Care Project Evaluation 2022 | Unable to assess |

### 2.2 Main findings by Intervention Type

#### 2.2.1 Discharge Planning, Teams and Pathways

Gonçalves-Bradley et al. (2022) performed a systematic review of RCTs to assess the effectiveness of **an individualised discharge plan for patients moving from hospital** as compared to routine discharge. Searches were completed in April 2021. Thirty-three RCTs were included of which 16 RCTs (**2 from the UK**) included data on older people. All patients in hospital (acute, rehabilitation or community) were included irrespective of age, gender or condition. For older people, authors concluded that a structured discharge plan that is tailored to the individual patient probably brings about a small reduction in the initial hospital length of stay and readmissions to hospital for older people with a medical condition and may slightly increase patient satisfaction with healthcare received. Specific conclusions were as follows:

*Mortality:* For older people with a medical condition (usually heart failure), from pooling 8 trials, it is **uncertain if discharge planning has an effect on mortality** at three- to nine-month follow-up (RR 1.05 (95% CI 0.85, 1.29) p=0.67).

*Hospital length of stay:* An analysis of 11 trials of older people found a mean difference in hospital length of stay that **just favoured discharge planning** (mean difference −0.73 (95% CI −1,33, −0.12) p=0.02. For two trials looking a hospital length of stay following surgery there was no difference; mean difference = −0.06 (−1.23, 1.11) p=0.92.

*Readmissions:* For older people, **discharge planning led to a relative reduction (from 17 trials) in readmissions to hospital** (average follow-up within three months; risk ratio (RR) 0.89 (95% CI 0.81, 0.97) p=0.01).

Quality appraisal rated the review as **moderate quality**.

Mabire et al. (2016) conducted a systematic review to examine the effectiveness of **discharge planning interventions that included at least one nurse** on health outcomes of older patients (≥65 years) going home from hospital. They also aimed to examine the impact of individual components of the interventions. The literature was searched from 2000 to 2015 and thirteen studies (10 RCTs, two pilot studies and one pre-post study) were identified for inclusion from across the world (none from the UK). A total of 3964 participants were included all of whom were aged over 60 years (median of 77 year or above in four studies), which should be noted is less than their inclusion criteria of 65 years or over.

Quality appraisal of the review revealed it to be of **critically low quality.** The results from the meta-analysis showed **no impact of nurse discharge planning on readmission rates or quality of life** (physical and mental domains), however it **significantly increased length of stay in hospital**. There was no significant heterogeneity between studies based on length of stay and omitting studies from the analysis did not change the conclusion.

Publication bias was possibly identified for readmission rates and removing studies from the analysis changed the conclusions. Subgroup analyses investigated heterogeneity between studies for readmission rates and suggested no impact of intervention type, provider type, measurement time, or age. However, the subgroup analysis suggested that in the USA nursing discharge planning reduces readmission. The authors note a great deal of variation in the interventions and that usual care was not always described. They suggest the strength of recommendations to be low or very low.

Jones et al. (2022) used **routinely collected data** across **England** between 2011 and 2016 to explore why **delayed transfers of care** (DTOCs) occur and how rates could be prevented or reduced. Alongside a range of data sources 31 discharge teams completed an online questionnaire. **Every extra home care provider** per 10km^2^ **decreased DTOCs** by 6.7– 8.0%, equivalent to 178–212 days per quarter for the average local authority, and a 1% rise in number of providers within 20km of a Middle Layer Super Output Area (MSOA) decreased DTOC by 0.17–0.18%, equivalent to two extra providers reducing DTOCs by 4.5–4.8 days per quarter. Urgent and Emergency Care (UEC) <u>Vanguard</u> partners (9/31 local authorities) had a 29.7% to 32.8% lower DTOCs compared to the other online survey sites. Of the case studies, 4/6 were UEC partners and on comparison **discharge planning appeared to be the key difference**. It was perceived that early planning gave more time to set up care packages and enable patients’ families to prepare for discharge. There were **no differences in the structure of discharge teams** or the **range of discharge pathways** available between the UEC vanguards and the other two case studies. It was concluded that physically co-locating social care and NHS discharge teams can assist with the visibility of teams as well as communication across disciplines. Clear discharge pathways are especially important where there is high ward staff turnover or use of agency nurses. Some DTOCs could be due to communication problems between organisational representatives. The authors could not explore how specific local discharge approaches and context can affect DTOC rates and discharge arrangements. The study was rated as **moderate quality**.

#### 2.2.2 Early Supported Discharge

##### Based on the definitions of each intervention (**Table 1**), this heading also includes interventions described as Early Discharge Hospital at Home, Discharge to Assess, Home Based Bridging and Home First

Davis et al. (2019) consulted frail older adults residing in **South Yorkshire** in **2014** about a recently adopted **discharge to assess** (D2A) service. The participants (n=27) were from black, Asian and minority ethnic communities, affluent and non-affluent areas and varied social circumstances. All participants **appreciated the need for hospital stays to be as short as possible** and if they were well enough they would prefer to be at home, even when their circumstances were not ideal. Participants considered discharge planning to be highly individualised and that those who were alone would need longer admissions. **Priorities** for discharge included **remaining independent** despite often feeling lonely at home; to **remain in hospital if needed**; and for services to ensure effective **communication with families**. The study was rated as moderate quality.

Fisher et al. (2020) used a **cohort study** design to investigate the effectiveness of **ESD** service models operating in real-world conditions. Using the national **stroke register of England** (Sentinel Stroke National Audit Programme; SSNAP) data was collected on 6260 patients between January 2016 and December 2016. Patients were clustered in 31 teams in regions within **England**. The majority of patients (91.9%) had a mild or moderate stroke (National Institutes of Health Stroke Scale <15). Although around 17% of patients were < 60 years of age, 64% were aged ≥70 years of age. Before their stroke, 4151 (66.3%) of patients were functionally independent (modified Rankin Scale=0). ESD service models were categorised with a 17-item score reflecting adoption of ESD consensus core components as outlined in an international consensus document and evidence-based postacute organisational audit criteria utilized by the SSNAP in the postacute audit. A range of ESD models had been adopted with total ESD consensus scores varying across the 31 teams from 5 to 15 (mean 10.6, SD 2.4). An increase in ESD consensus score was associated with a more responsive ESD service, this association appeared to be driven by having **more core team members** meeting or exceeding recommended whole time equivalent level per 100 patients with stroke (a 1-unit increase in the consensus score was significantly associated with a 47% reduction in the odds of the ESD team seeing the patient after 1 day or more following hospital discharge [95% CI, 14%–67%], p=0.01). Thus, improvements in the team components (see *Extract* below) were directly correlated with improved hospital to home care transition. The study was well reported and rated as **high quality**.

**Table 1.**
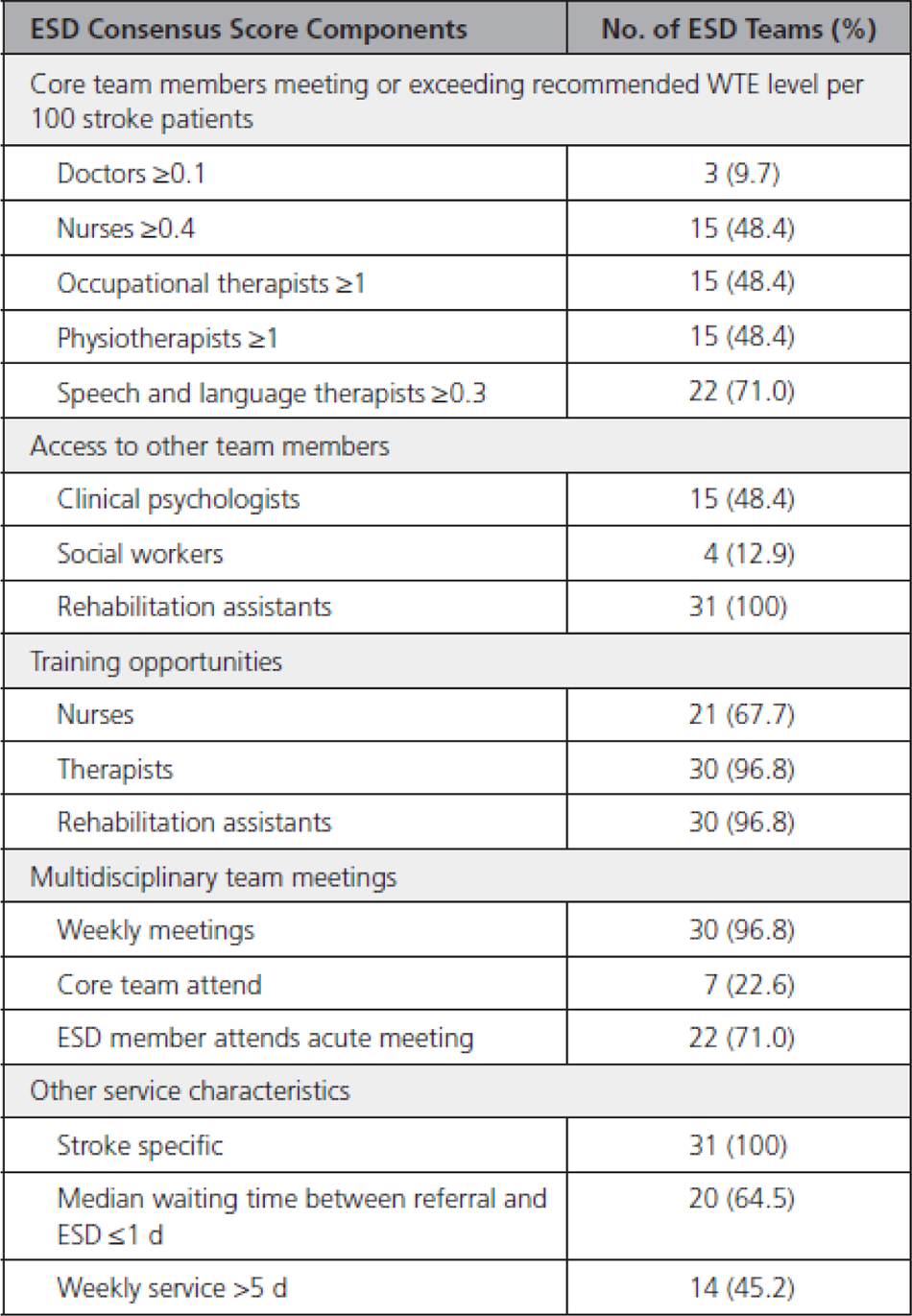
ESD Consensus Score Components Across 31 ESD Teams.

Gonçalves-Bradley et al. (2017) performed a systematic review of RCTs to determine the effectiveness and cost of managing patients with **Early Discharge Hospital at Home** compared with inpatient hospital care. Searches were completed in January 2017. 32 RCTs were included (**16 in the UK**) of which 16 RCTs (**8 in the UK**) provided data for older people aged 65+. Authors noted that descriptions of Early Discharge Hospital at Home varied but **in the UK** the focus is usually on the provision of personal, nurse-led care, building on the existing structure of primary care. Authors concluded that the review provided low- to moderate-certainty evidence that hospital at home does not adversely affect mortality, hospital readmission, or functional status. The following conclusions were drawn based on the findings for people aged 65+:

*Mortality:* Early discharge hospital at home probably makes **little or no difference in mortality** to older people with a mix of conditions, or COPD. Twelve trials reported data for mortality at three to six months follow-up for older people with a mix of conditions, and data were pooled from eight of them (RR 1.07 (95% CI 0.76, 1.49) p=0.71) and from five trials recruiting patients with COPD (RR 0.53 (0.25, 1.12) p=0.10).

*Hospital readmission:* Early discharge hospital at home probably **increases the risk of readmissions** for older people with a with a mix of conditions and **may decrease the risk of readmissions for people with COPD**. Pooling data for nine trials recruiting older people with a mix of conditions, median follow-up of three months (RR 1.25 (95% CI 0.98, 1.58; p=0.07), and five trials for participants with COPD with two to three months follow-up (RR 0.86 (0.66,1.13) p=0.29).

*Functional status*: Based on the findings of four trials, Early discharge hospital at home probably makes **little or no difference to functional status** (Barthel Index; mean difference 0.34 (95% CI −0.18, 0.86) p=0.20). There was substantial heterogeneity between trials.

*Patient reported outcomes:* There was **little or no difference on a range of patient reported outcomes** between groups for older people with a mix of conditions and patients with COPD.

*Institutional care:* Based on two trials with one year follow up and one with 6 months the RR for **institutional care at one year follow up (just significantly) in favour of early discharge**: Risk Ratio 0.69 (95% CI 0.48, 0.99); p=0.04.

*Patient satisfaction:* Of six trials reporting patient satisfaction, two reported increased levels of satisfaction for those allocated to early discharge hospital at home and four trials reported little or no difference.

*Caregiver outcomes:* Five trials measured caregiver outcomes, including strain and general health. Three reported little or no difference (Cunliffe 2004; Shepperd 1998; Utens 2012), while two found less caregiver strain in early discharge hospital at home (Harris 2005; Tibaldi 2013). Three trials reported that early discharge hospital at home may increase carer satisfaction (Harris 2005; Ojoo 2002; Utens 2012), and two reported little or no difference (Caplan 2006; Shepperd 1998).

*Staff views:* One trial reported that staff perceived that providing care in the patients’ homes facilitated participation in rehabilitation, that the service was better staffed than the usual discharge services provided, and that rehabilitation services were coordinated with social care (Cunliffe 2004; results not tabulated); and a second trial reported little or no difference in general practitioners’ level of satisfaction (Caplan 2006).

*Length of hospital stay:* Combining data for four trials found that early discharge hospital at home probably **reduces hospital length of stay** (mean difference −6.76 days (95% CI − 10.60, −2.92) p=0.0006. There was substantial heterogeneity. In contrast to the findings above, pooling data from three trials that reported both length of stay in hospital and hospital at home suggested that early discharge hospital at home may increase the number of days of health care received (mean difference 6.43 (2.84,10.03) p=0.0005).

*Use of healthcare resources and cost:* Seven trials reported the costs associated with the intervention. There were varied outcomes for different settings and populations. Hospital at home may be more, less or the equivalent expense as in-hospital care.

This is a vast review with multiple analyses, but quality appraisal rated it as **critically low quality**.

Hywel Dda University Health Board in **2022** conducted a **case study** of **Home Based Bridging Care** and provided interim findings (Home Based Bridging Care Project Evaluation 2022). However, it should be noted that one of the main evaluation measures, **staff recruitment**, failed to reach target levels and prevented a full assessment of the impact of the workforce on outcomes. At the end of January 2022, 16.60 WTE had been recruited against a target of 60 WTE with all bar one person having commenced in post.

Pembrokeshire was the only county in the Health Board area to have **deployed staff specifically into a bridging service** and this because there was an existing team already supporting this function, early indications suggest that it has supported growth of the caseload and activity. The impact on **discharge delay** found that the average number of “**lost days**” between the date ‘ready to leave’ and discharged is **7 days** across the Health Board with some **considerable variation between sites** and the range is significant from 0 to 117 days. A quality rating is not presented as there is no appropriate quality appraisal tool to appraise this level of evidence when the population comprises a region.

Langhorne et al. (2017) conducted a systematic review examining the impact of **services offering a policy of early discharge with rehabilitation (early discharge support)** to people in hospital with a **stroke** in terms of speed of return home, carer and patient outcomes, acceptability to patients and carers and resource implications. Searches were completed up to Dec 2016 to Mar 2017 (depending on source) for RCTs and 17 trials from around the world were included in the review. Average ages of participants ranged from 60 to 80 years; however sub-group analyses were conducted to examine the impact of age by splitting the data into two groups of < 75 and > 75 years of age. Nine trials were included in the subgroup analyses, **four of which were from the UK**. Sample sizes of these nine trials ranged from 23 to 331. Quality appraisal of the review revealed it to be **critically low quality**. The overall analyses (participants of all ages) showed **reduced death or dependency** (median 6 months; range 3 to 12 months; OR 0.80, 95% CI 0.67, 0.95, p=0.01; moderate-grade evidence) in the intervention groups as well as **reduction in length of stay** (mean difference −5.5, 95% CI −3 to −8 days, p<0.0001; moderate-grade evidence). The subgroup analyses for death or dependency (main outcome) and length of stay showed that **age was not associated with the effect of early supported discharge** (results similar for <75 and >75 years). The authors note that their inclusion criteria for interventions was purposefully broad.

Williams et al. (2022) performed a systematic review of RCTs to explore the evidence for **early supported discharge in older adults** hospitalised for acute medical care. Searches were undertaken in January 2021. Five studies were included (one from UK) of MDT led interventions based in participants homes (average participant age ranged from 79.8 to 83.8) of varying duration and intensity. Length of hospital stay was the primary outcome investigated. Implementation of early supported discharge was associated with a **significant reduction in length of stay** (data from 4/5 included studies, Mean difference = −6.04, CI −9.76 to −2.32, p=0.001). Results on **intervention costs were variable** (one study found the intervention to be associated with higher costs, and two found the care of the control group was more costly). No statistically significant results favouring the ESD interventions were identified in relation to mortality, function, quality of life, hospital readmissions or cognition. Confidence in the review findings was rated as **critically low**.

##### 2.2.2.1 Summary of Evidence for Early Supported Discharge and Hospital Length of Stay

Three systematic reviews were included that evaluated the effect of early supported discharge on **hospital length of stay** (Gonçalves-Bradley et al. 2017; Langhorne et al. 2017; Williams et al. 2022). The outcome effect was **consistent** across the 3 systematic reviews in that early supported discharge was associated with a **reduction in length of stay**. The reviews estimated the mean reductions in stay 6.8 days (Gonçalves-Bradley et al. 2017), 5.5 days (Langhorne et al. 2017) and 6 days (Williams et al. 2022). The 3 systematic reviews were rated as either **low or critically low quality**.

From within these three systematic reviews (Gonçalves-Bradley et al. 2017; Langhorne et al. 2017; Williams et al. 2022), four UK primary studies (all RCTs) that were published since 2000 were identified, suggesting a lack of recent UK evidence. The studies were not quality appraised, but data about length of hospital stay (and readmission) were extracted (section 6.2.3). Two studies involved MDTs (Cunliffe et al. 2004; Donnelly et al. 2004) and two involved respiratory nurses only (Cotton et al. 2000; Ojoo et al. 2002). Three studies concerned patients with specific conditions (Cotton et al. 2000; Donnelly et al. 2004; Ojoo et al. 2002). Regarding **hospital stay data** the results did **not consistently present significant differences** in favour of the intervention; two studies indicated reduced hospital stay in the intervention groups (Cotton et al. 2000; Cunliffe et al. 2004), but two studies showed no significant differences (Donnelly et al. 2004; Ojoo et al. 2002). Regarding **readmission** none of the studies found any significant differences between groups.

#### 2.2.3 Intermediate Care

Elston et al. (2022) used a **controlled before and after** study design to compare the **Enhanced Intermediate Care** (EIC) service in one locality of the **Torbay and South Devon NHS Foundation Trust** to **Intermediate Care** services in 4 other localities within the same trust. Performance data was collected between April 2015 and March 2018. The EIC area showed statistically significant increase in EIC referrals to 11.6% (95%CI: 10.8%–12.4%); **more people being cared for at home** (10.5%, 95%CI: 9.8%–11.2%), **shorter episode lengths** (9.0 days, CI 95%: 7.6–10.4 days) and **lower bed-day rates** in ≥70 year-olds (0.17, 95%CI: 0.179–0.161). The study was rated as **moderate quality**.

Levin and Crighton (2019) used an **interrupted times series** to measure the effect of **Intermediate Care (IC) and a 72-hour discharge target** on days delayed for patients aged ≥ 75 years of age. Data was collected between January 2013 and June 2016 comparing **Glasgow City** before and after onset of IC with **Inverclyde and West Dunbartonshire** as a control. Approximately 63% of the resident population were female but a greater number in Glasgow were in the most deprived quintile. In Glasgow City**, IC combined with the 72-hour discharge** was associated with a **step reduction in the rate of bed days delayed per population** [−15.20 (95% CI −17.52 to –12.88)] and a **reduced upward trend** in rate of bed days delayed in the longer term [−0.29 (95% CI −0.55 to –0.02)]. **Rate of days delayed continued to increase over time, although at a slower rate than if IC had not been implemented.** The authors noted several limitations: the **observed effect** might be an **underestimate** due to residents from the control area being able to attend the same hospitals as residents of Glasgow City; introduction of IC may impact discharge behaviour and also require ‘bedding in’ and further refinements. The study was rated as **moderate quality**.

The National Development Team for Inclusion (2019) conducted a **pilot** study using **mixed methods** to explore the impact of the intermediate care **Shared Lives model** for people who are ready to leave hospital, but unable to return home with the aim to **reduce the length of hospital stay**, and or to prevent the need for admission to hospital or long-term residential care by providing alternative support for a limited period of time. Only small numbers were involved therefore quantitative data to measure impact on length of stay is not available. Of 58 referrals, 31 patients were successfully placed of which 10 eventually returned home and 11 entered a long-term Shared Lives placement. Of those who were placed most were **<65 years of age**. The **numbers of people** discharged from hospital and supported via the Shared Lives Intermediate Care programme are **too low** for the pilot to have made any **noticeable impact** on the local health and social care system. Pilot sites found it difficult to access health teams and health funded placements failed to materialise. It was noted that it was a challenge to get health professionals to understand and trust the model. New contacts were made with mental health teams who referred the largest number of people. People with complex and multiple needs benefitted such as those with mental ill health or inappropriate housing circumstances. Pilot sites showed flexibility, adapting systems and processes. Shared Lives carers were mostly existing carers and were limited in geographical spread. People in Shared Lives arrangements said that having ‘a life’ and feeling connected is important to their health and happiness. This was a pilot project and only small numbers involved therefore quantitative data to measure impact on length of stay is not available. Therefore, only the qualitative methods have been quality appraised. The study was rated as **low quality**.

#### 2.2.4 Multidisciplinary rehabilitation (post hip fracture)

Handoll et al. (2021) completed a systematic review to examine the effects of **multidisciplinary rehabilitation** within inpatient or **ambulatory care settings on older people who had a hip fracture.** The ambulatory care setting covered home (including nursing homes), outpatient departments and day hospitals. Interventions that were predominantly conducted within ambulatory care settings (including those that were mixed community and hospital settings) were analysed separately to those conducted inpatient settings. This was an update to a review, so search dates were from 2009 (previous review) up to Nov 2019 (trial registers) or Oct 2020. Twenty-eight randomised trials from around the world were included in the review, with seven **(one from the UK)** being included in the extracted section focusing on interventions in ambulatory care settings. The review did not specify ≥65 years in the inclusion criteria but they used exclusion criteria to minimise the impact of younger participants. Of note, only one study (Singh et al., 2012) in the extracted section had a lower age limit of <65 years and this study had a mean age of almost 80 years. Sample sizes of the seven trials ranged from 53 to 240. Quality appraisal of the review revealed it to be of **moderate quality**. Results were pooled from three studies that compared **supported discharge and multidisciplinary home rehabilitation** compared to usual care for those people who mainly lived at home. There was very low certainty in the evidence of **little to no difference between groups** in **‘poor outcome’** (death or move to a higher level of care or inability to walk), quality of life, mortality, independence in daily activities, permanently moving to higher care and inability to walk (most measured at 12 months, number of studies varied by outcome.

One study compared **supported discharge and multidisciplinary home rehabilitation** compared to usual care for nursing home residents. Low certainty evidence of **no or minimal group differences** were reported for **‘poor outcome’** (dead or unable to walk at 12 months) and **mortality**. Very low certainty evidence of **no difference between groups for dependency, quality of life, inability to walk or pain** were reported.

Another study examined **intensive with less intensive multidisciplinary home-based rehabilitation** (from the UK) and reported have very low certainty evidence for **no differences between groups on any outcomes**.

A study which examined **multidisciplinary care (including progressive resistance training for a year)** compared to usual care included very low certainty evidence of **marginally favouring the intervention**.

The authors of the review note considerable variation in the characteristics of the interventions and usual care. They also caution that the results should be interpreted in the context of heterogeneity in interventions, populations and measures.

#### 2.2.5 Transitional care/continuity of care

Baxter et al. (2020) explored **staff perceptions** in the **North of England** between September 2017 and May 2018 of how high performing general practice and hospital specialty teams deliver **safe transitional care** to older people as they transition from **hospital to home**. Staff (n=157) were from a **range of health disciplines but not social care**. In addition to the staff focus groups and interviews, 9 discharge meetings were observed. **Three themes** emerged of how safe transitions were facilitated: **getting to know the patient**; building r**elationships** within and across **teams** and **bridging gaps** within the **system**. **Transitions appeared safest** when **all 3 themes were in place** but this was not always possible particularly across settings. It was **easier to overcome these challenges** for **patients with complex transitional care needs.** The study was well reported and deemed to be **high quality** with no substantial areas of concern. Although the authors suggest that there might be bias in the participant responses who may not be representative and the researchers may have had biases that influenced the work.

Facchinetti et al. (2020) performed a systematic review of RCTs to evaluate the effectiveness of **continuity of care in older people (≥ 65 years) with one or more chronic diseases** in reducing short- and long-term hospital readmission after hospital discharge as compared to routine care. Searches were completed in January 2019. 36 RCTs were included (**3 in the UK**) exploring continuity of care interventions focusing on the connection and coordination between patients and providers across time and settings and classified in informational, management, and relational continuity contexts. Outcome measures explored in meta-analyses (incorporating 30 RCTs) were hospital readmissions in time sections up to 12+ months post discharge. 8,920 patients were included; 53% with chronic heart failure, 10% chronic obstructive pulmonary disease (COPD), 7% COPD and heart failure, 3% chronic lung disease. No other demographic details were provided. At **one month from discharge, the continuity interventions were associated with lower readmission rates** (12.9% in the experimental group and 16% in the control group; Relative Risk [RR], 0.84 [95% CI, 0.71-0.99] p=0.04). **From 1 to 3 months, readmission rates were lower in the experimental group** (21.9%) versus the control group (29.8%; RR 0.74 [95% CI, 0.65-0.84] p<0.00001). A subgroup analysis showed that this positive effect was stronger when the interventions addressed all of the continuity dimensions (patient provider relationship, transfer and use of information, management continuity). **At 3-6 months this impact became inconclusive** with moderate/high statistical heterogeneity. Although not included by authors in the abstract, **data for 6-12 months suggested benefit** (RR 0.84 (0.74, 0.95) p=0.007). Quality appraisal of the review rated it as **critically low quality**.

Lee et al. (2022) completed a systematic review and meta-analysis of the **effect of transitional care, from hospital to home**, on the health of frail older adults (≥65 years). Searches were completed up to August 2021 and identified 21 included studies which reported on 14 trials, two of which were from the UK (and others from across the world). Only RCTs were eligible for inclusion in the review. A total of 5776 participants were included (sample sizes ranged from 128 to 2353), aged between 77.0 and 85.7 years. The interventions varied in terms of their setting, components, healthcare providers and duration. Outcome measures and assessment times also varied between studies. Quality appraisal of the review revealed it to be **critically low quality.** Results from the meta-analysis showed that transitional care interventions **reduced re-admission at six months** but not at other time points (earlier and/or later). However, the results suggest heterogeneity. No impact on mortality was shown, neither was there an impact of the intervention on health-related quality of life, but similar to readmission at six months, heterogeneity was indicated. There were mixed results for physical functioning, cost and satisfaction, but all of the studies examining self-related health or satisfaction reported that the interventions had a positive effect. The authors observed that higher intensity interventions appeared to be more effective, though they did not classify interventions based on this within the review. The authors conclude that the **effectiveness of transitional care interventions for frail older adults is unclear**.

Meulenbroeks et al. (2021) performed a systematic review of RCTs and non-RCTs to assess whether **transitional care programs that integrate caregivers** provide better value care than routine care for **people with geriatric syndrome** (aged 65+ and displaying moderate to severe geriatric traits as measured by validated tool). The intervention compared caregiver inclusive transitional care from acute to community settings with routine care. 23 studies in high income countries were included **(two in the UK)**; 14 RCTs and 9 NRCTs, 90% rated as with high or critical quality concerns. 16,657 patients were included in all, with an average age of 77.8. A narrative analysis only was provided. Authors found that **consistently positive results occurred for patient and caregiver satisfaction**. **Cost tended to increase with caregiver inclusive practices. Most studies found no difference in population health outcomes.** They concluded that there was insufficient evidence on healthcare professional experience and insufficient evidence to determine whether caregiver inclusive transitions of care provide *better value* care than routine care. Studies that rigorously implement and evaluate caregiver inclusive care models are urgently required to inform future policy. Quality appraisal rated the review as **critically low quality**.

Rasmussen et al. (2021) investigated the impact of **transitional care interventions with both pre- and post-discharge components** on readmission of older medical patients. They conducted a systematic review searching the literature from January 2008 to August 2019. Eleven studies of differing design (five RCTs, four non-randomised controlled trials and two pre-post cohort studies) were included, with **one of these being from the UK** and others from across the world. A total of around 24500 participants were included (sample sizes ranged from 41 to 19157). Studies were included if participants were aged 65 years or above, or the mean study population was over 75 years. Mean ages were reported to be ∼78 (range 74.9 to 83.6) and ∼79 (range 75.2 to 84.5) years for intervention and control groups respectively. However, data within the table of study details reported that the age range for one study had a lower limit of 60 years. Interventions differed most on the pre-discharge phase components; bridging and post-discharge components were fairly similar. Quality appraisal of the review revealed it to be of **low quality.** Results of the review suggested that the **majority of the results (22/29) measuring re-admission showed lower rates in the intervention groups** compared with controls. However only five of the results were statistically significant. Subgroup analyses showed a greater impact on readmission rates 1) for **high risk patients**, 2) for **higher intensity interventions**, 3) for support **of a month or more**, 4) in **non-European** studies (indeed all statistically significant results were from non-European countries), 5) when measured within a month of discharge (though statistically significant results were all of assessments between one and three months). The authors note that the **certainty of the evidence is low** using GRADE. They also note that when contacting researchers in the field they reported either struggling to or not publishing negative findings thus negative findings may be under-represented in the review.

Tomlinson et al. (2020) performed a systematic review and meta-analysis exploring interventions that focused on **supporting medication continuity** among older people and their impact on hospital readmissions. RCTs were included where participants had a mean age of ≥65 years. Searches were undertaken for the period 2013 to September 2019. Of the 24 included studies, 15 were relevant to our review (**one was set in UK**) as they either began in hospital and continued in the community (9) or were initiated in the community (6). There was significant variation in the interventions identified including populations, care settings, intervention components, intensity and duration. Community interventions were generally pharmacist led, while those which began in hospital were also delivered by nurses or a multidisciplinary team. **None** of the interventions that **commenced in the community** showed a statistically significant reduction in hospital readmission (all were considered high quality). The included UK study (Holland et al., 2005) showed a 30% higher readmission rate in the intervention arm. **Five of the nine interventions commenced in hospital**, which continued between 7 and 180 days post-discharge showed a **reduction in hospital readmissions** (though 4 were considered to have great concerns in quality). Higher intensity multi-component interventions were more likely to be successful. Meta-analysis across 19 of the included studies identified **self-management education** or coaching, **telephone follow-up** and **medication reconciliation** as the activities associated with reduced readmissions although the authors cautioned that **firm conclusions could not be drawn** about causality. Quality appraisal of the review rated it as **moderate**.

##### 2.2.5.1 Summary of body of evidence for Transitional care/Continuity of care and Readmission

Four systematic reviews were included that evaluated the effect of Transitional care/Continuity of care **on readmission** (Facchinetti et al. 2020; Lee et al. 2022; Rasmussen et al. 2021; Tomlinson et al. 2020). The **overall outcome effect** tended to demonstrate a **reduction** in readmissions. Facchinetti et al. (2020) found that at 1 and 1-3 months post-discharge interventions were associated with **lower** readmission rates. Lee et al. (2022) found that **transitional care interventions reduced re-admission at six months** but not at other time points. Rasmussen et al. (2021) found that the majority of the included studies (22/29) measuring re-admission **showed lower rates in the intervention groups** compared with controls. However only **5** of the results were **statistically significant**. Tomlinson et al. (2020) found in 5/9 studies that there was a reduction in hospital readmissions. The 4 systematic reviews were rated as either **low or critically low quality**.

From these four systematic reviews (Facchinetti et al. 2020; Lee et al. 2022; Rasmussen et al. 2021; Tomlinson et al. 2020), five UK primary studies (RCTs) since 2000 were identified. They were not quality appraised but data about readmission (and hospital stay) were extracted (section 6.2.4). There was limited recent evidence (Sahota et al. 2017). Two studies involved specialist nurses only (Blue et al. 2001; Cleland et al. 2005), one study involved pharmacists only (Holland et al. 2005), one study involved specialist geriatric medical management but the professionals were not specified (Edmans et al. 2013) and one study involved a MDT (Sahota et al. 2017). Two studies concerned patients with specific conditions (Blue et al. 2001; Cleland et al. 2005). Regarding **readmission**, the results showed **no consistent pattern**. Two studies indicated more readmissions at 6 months (Holland et al. 2005) or hospital presentations at 90 days (Edmans et al. 2013, though no significant difference in days spent at home) in the intervention group but one study reported fewer readmissions in the intervention group at 12 months (Blue et al. 2001). Furthermore, one study reported no significant difference in readmission at 28 days (Sahota et al. 2017) and one did not analyse group differences (Cleland et al. 2005). Regarding hospital length of stay of the two studies reporting this outcome Blue et al. (2001) noted a reduction in length of stay while Cleland et al. (2005) did not identify any significant difference. A cost-effectiveness analysis of an included RCT (Tanajewski 2015) was also identified from one of the studies (Edmans et al. 2013). Tanajewski (2015) concluded that the specialist geriatric medical intervention for frail older people discharged from acute medical care was not cost-effective.

Three themes emerged from staff perceptions reported by Baxter et al. (2020; UK study) with regard to how safe transitions should be facilitated, these were: **getting to know the patient**; building r**elationships** within and across **teams** and **bridging gaps** within the **system**.

#### 2.2.6 Bottom line results

- Moderate volume of evidence, 11/19 studies are systematic reviews
- None of the systematic reviews exclusively include primary studies conducted in the UK
- 16/19 of the included studies were published as journal articles
- The 19 studies evaluated 5 different intervention areas and a range of outcomes
- None of the systematic reviews were rated as high quality
- 1 primary quantitative study was rated as high quality and the other 3 were rated moderate.
- The mixed methods study was rated low quality
- 1 qualitative study was rated high quality and the other moderate
- 1 case study was included of Home Based Bridging Care in the Hywel Dda University Health Board region
- The body of evidence for Early Supported Discharge and hospital length of stay is consistent and indicates that early supported discharge reduces length of stay. 4 recent UK studies however, identified from the included systematic reviews, did not consistently present significant effects of the intervention on length of stay.
- The body of evidence for Transitional care/Continuity of care and readmission is fairly consistent and tended to demonstrate a reduction in readmissions. 5 recent UK studies however, identified from the recent systematic reviews, have not confirmed this finding.

## 3. DISCUSSION

### 3.1 Summary of the findings

This **rapid review identified** and included a moderate volume of evidence (**n=19**). The search for this review focused on identifying systematic reviews, of which 11 were included and a search for more recent primary studies specifically conducted in the UK.

Although there was a moderate volume of **evidence** it was **spread across several different interventions** which were evaluated by a **variety of outcome measures**. All the **interventions** evaluated were **multi-disciplinary**.

Two interventions that had been evaluated in the most studies with the same outcome were: Early Supported Discharge (ESD) with hospital length of stay and Transitional care/Continuity of care with readmission. The body of evidence for ESD and hospital length of stay indicted that **ESD reduces hospital length of stay.** Also, the body of evidence for **Transitional care/Continuity of care** and readmission is fairly consistent and tended to demonstrate a **reduction in readmissions**. However, it should be noted that these findings are derived from systematic reviews and likely that there is overlap and double counting of studies. Also the descriptions and components of each of the interventions is likely to vary, particularly as the systematic reviews are not limited to UK primary studies. The recent (2000 onwards) UK studies from these two interventions were explored in more detail and showed that 1) length of stay was reduced in some but not all studies of ESD and 2) findings about readmission following transitional care/continuity of care were inconsistent.

### 3.2 Limitations of the available evidence

None of the studies identified focused specifically on community care alone in relation to hospital discharge.

Geographical location is likely to add to the heterogeneity in intervention designs and outcomes. None of the systematic reviews included in this review included only studies from the UK and there are relatively few UK studies within the international systematic reviews. All systematic reviews apart from Mabire et al. (2016) included at least one UK study, and for one review (Goncalves-Bradley 2017) half of the studies were from the UK.

Where systematic reviews covered roughly the same intervention and outcomes there is likely to be overlap in the included primary studies.

Significant differences in the interventions delivered (including the health care professionals involved, components, intensity, duration) make it difficult to provide clear recommendations for optimum service design.

In terms of quality, none of the systematic reviews were of high quality and only 1 of the 4 quantitative studies were rated as high quality.

### 3.3 Implications for policy and practice

It is difficult to draw firm conclusions due to the limited evidence from a UK setting, and low quality of included studies.

The global evidence has identified the potential for early supported discharge and transitional care interventions to impact on length of hospital stay and readmissions. This has not been confirmed by the small amount of recent UK evidence and further research is indicated within UK settings.

There is insufficient information from the research studies included in this review to propose an optimum service design but the evidence does appear to suggest that interventions that are more comprehensive (covering a range of different components), and more intensive are more likely to be effective.

There are some recent publications looking at good practice in the Discharge to Assess/Home First model that has been adopted in Wales. These are not research publications, so fall outside this review, but both include examples of service models in Wales^3^ and England^4^. There is no consensus guidance on a work-force model within the Welsh Government document^3^ but the English guidance^4^ provides expert advice on the discharge process, including roles, structures and responsibilities. These include the following.

- An executive lead to provide strategic oversight of the discharge process.
- A single coordinator to secure safe and timely discharge on the appropriate pathway for each individual.
- A transfer of care hub associated with each acute hospital to link services (health, social care, housing, voluntary sector) together along with support for unpaid carers. Hubs to include a case manager to coordinate care and support the individual.
- A hospital-based multidisciplinary team (including a hospital based social worker) to describe – with input from the person and their unpaid carer, advocate, or relevant community-based professionals – the needs that require support after discharge before an assessment of their long-term needs.
- Direct links to primary care providers.

In December 2021, NHS England established a National Discharge Taskforce (chaired by Sarah-Jane March, chief executive of Birmingham Women’s and Children’s Hospital Foundation Trust) to develop best practice in improving hospital discharge.

### 3.4 Strengths and limitations of this Rapid Review

This review was conducted rapidly to inform policy and decision makers. The review consists of 11 systematic reviews which comprise of internationally conducted primary studies which included at least one UK study, except for one review (Mabire et al. 2016) which did not include any studies from the UK. Though it is a strength to identify 11 systematic reviews there is likely to be overlap of included studies between the systematic reviews. As most of the included studies were conducted across the globe the findings might be limited in their generalisability to UK populations.

The 8 included primary studies were based in the UK and were mostly appropriately conducted with some areas of concern. These studies were not included in the 11 identified systematic reviews. Nine recent primary UK studies from the two bodies of evidence on Early Supported Discharge and hospital length of stay and on Transitional care/Continuity of care and readmission were unpicked from the systematic reviews to explore the consistency of results and outline intervention components. These additional primary studies were not quality appraised but, overall, evidence from 17 UK primary studies is included in the review.

It should be noted that given the lack of time the literature search was focused on the overarching aim of timely discharge. In addition to the 4 bibliographic databases, the grey literature search consisted of organisation websites and reports identified by the review team or provided by Stakeholders.

In conducting this rapid review, quality appraisal and data extraction of each study was undertaken by different reviewers and not independently in duplicate or checked for accuracy and consistency.

## Abbreviations

Acronym Full: Description

AMSTAR: Assessment of Multiple Systematic Reviews

BAME: Black, Asian and Minority Ethnic

CI: Confidence Intervals

COPD: Chronic obstructive pulmonary disease

D2A: Discharge to Assess

DTOC: Delayed Transfers of Care

EIC: Enhanced Intermediate Care

ESD: Early Supported Discharge

IC: Intermediate Care

IWD: Inverclyde and West Dunbartonshire

MDT: Multi-Disciplinary Team

MSOA: Middle Layer Super Output Area

NDTI: National Development Team for Inclusion

NRCT: Non-Randomised Controlled Trial

RCT: Randomised Controlled Trial

RoB: Risk of Bias

RR: Relative Risk

SD: Standard Deviation

SSANP: Sentinel Stroke National Audit Programme

UEC: Urgent and Emergency Care

UK: United Kingdom

## Data Availability

n/a

## 5. RAPID REVIEW METHODS

### 5.1 Eligibility criteria

The JBI rapid review inclusion criteria framework was used to define the criteria for this rapid scoping review, Participants, Concept, Context.

|  | **Include** | **Exclude** | **Notes** |
| --- | --- | --- | --- |
| **Population** | Hospitalised older adults (≥65 years of age) who are medically fit and waiting discharge to either the community or residential care. Includes all reasons for hospital admission. |  | Medically fit also known as 'medically fit for discharge' or 'clinically optimised' |
| **Intervention** | Workforce model to increase capacity for community care.<br>To include all aspects of care, including occupational therapy, speech therapy, physiotherapy<br>To include all possible scenarios, e.g. integrated care, MDTs, upskilling or reskilling or redeployment of existing staff, realignment of teams, patient reablement, overseas recruitment. | Unpaid carers or volunteers unless part of MDT approach.<br>In-hospital interventions such as those to reduce functional decline or medication counselling etc to reduce length of stay or reduce readmissions. |  |
| **Comparison** | Usual care |  |  |
| **Outcomes** | Numbers/rates of staff or workforce in community<br>Hospital length of stay<br>Number/rates of delayed transfers of care<br>Readmissions (<2 weeks)<br>Mortality<br>Cost analyses, within UK setting<br>Sustainability metrics |  | Other outcomes will be considered if noted to be a primary outcome measure in identified studies. |
| **Other considerations** | Prioritize SRs, supplemented with UK |  | To be confirmed after further scoping. |
|  | primary comparative studies<br>SRs from 2016-2022, need to have followed SR process.<br>Primary studies from 2019-2022 |  | Due to time constraints only limited searching of grey literature from key organisations. |

### 5.2 Literature search

This review was conducted according to a priori protocol. An initial scoping search was conducted to identify relevant systematic reviews published between 2020-2022. Following this scoping exercise, a search was completed to identify relevant primary research that was both published and unpublished from a wide-ranging set of resources. Known literature was provided by stakeholders and this was also checked for eligibility and included or used as a source of specific relevant evidence. For details of all the resources searched, please refer to Appendix 1.

### 5.3 Database search

The systematic review scoping search was conducted in Epistemonikos, details of the search strategy are found in Appendix 2.

Following this scoping exercise, a comprehensive search was designed in Medline [Appendix 2] to identify relevant primary studies and then translated across 3 other databases (HMIC, SCOPUS and Social Policy & Practice). It used a combination of text words and medical subject headings. The results of all the database searches can be found in Appendix 1.

### 5.4 Supplementary search

The grey literature search consisted of organisation websites and reports identified by the review team or provided by Stakeholders. For searching grey literature resources a broad search was conducted using word variations of the terms: “discharge” “rehabilitation” domiciliary care” “nursing home” “home base care” “organised home care” “mobile team” “hospital to home care” “primary care” “community care” [results reported in Appendix 1].

### 5.5 Reference Management

Database searches were imported into Endnote 20 and deduplicated. Grey literature search results were added to an Excel spreadsheet and cross-checked against the Endnote library.

### 5.6 Study selection process

Evidence selection from the database searches was conducted by an individual reviewer(s). Eligibility criteria were used to assess the titles and abstracts and then full text of all sources identified by the search. Grey literature reports were identified by individual reviewers and checked for eligibility. Where one reviewer was uncertain as to inclusion it was checked by a second reviewer.

### 5.7 Data extraction

Data were extracted from studies and reports into an Excel form to capture key information such as participants, indicators investigated, evidence type, data collection or literature search dates. Data extraction was carried out by individual reviewers.

### 5.8 Quality appraisal

Quality appraisal of the 19 included studies was completed to assess the trustworthiness, relevance and results reported. It was completed by a single reviewer using one of the following validated quality appraisal tools:

- AMSTAR 2: Critical appraisal tool for systematic reviews that include randomised or non-randomised studies of healthcare interventions, or both
- CASP Qualitative Studies Checklist
- JBI Critical Appraisal Checklist for analytical cross sectional studies
- JBI Critical Appraisal Checklist for quasi-experimental studies

### 5.9 UK primary studies identified within systematic reviews

For the two bodies of evidence identified, the relevant systematic reviews were examined to identify UK primary studies published since 2000. Data extraction (including study type, intervention components and readmission and length of stay findings) from these additional primary studies was completed by individual reviewers.

### 5.10 Synthesis

A narrative approach was used, including tables detailing the extracted data (authors (year), country, title, study details, population and settings, key findings and observations/notes), to provide descriptive summaries of the selected studies to the reader. This type of analysis is recommended for rapid reviews (Grant & Booth 2009).

## 6. EVIDENCE

### 6.1 Study selection flow chart

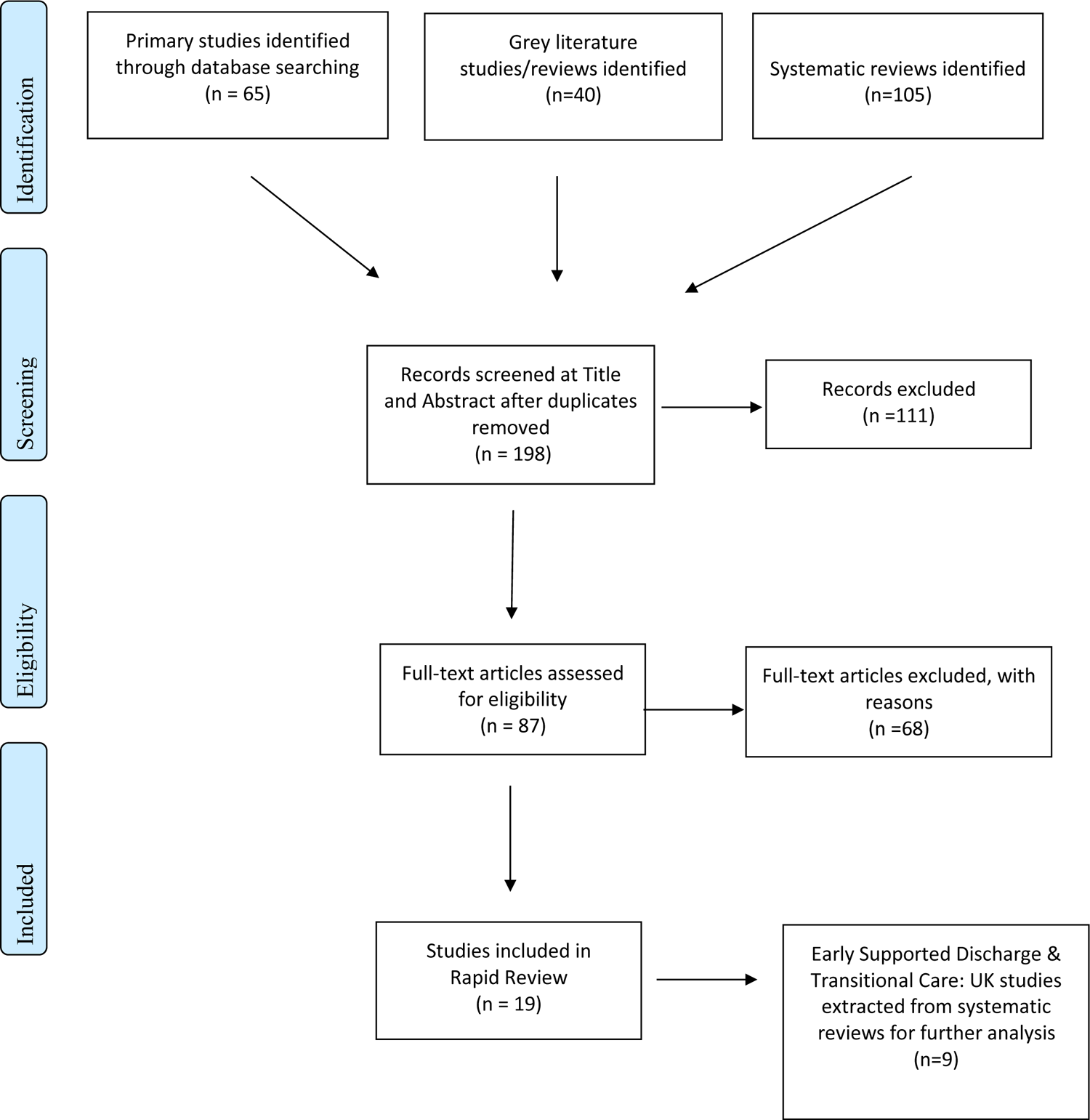

### 6.2 Summary of included research

#### 6.2.1 Secondary Research (Table 2)

| Citation | Review details | Included studies | Quality | Findings and observations/notes |
| --- | --- | --- | --- | --- |
| **Facchinetti et al. 2020**<br>Continuity of care interventions for preventing hospital readmission of older people with chronic diseases: A meta-analysis. International journal of nursing studies. 101: 103396. doi: 10.1016/j.ijnurstu.2019.103396 | **Review aim:** To evaluate the effectiveness of continuity of care interventions in older people with chronic diseases in reducing short and long term hospital readmission after hospital discharge<br>**Population:** older patients (≥ 65 years) diagnosed with one or more chronic diseases<br>**Intervention:** Continuity of care interventions provided by any healthcare professional during and after <u>hospital discharge</u>. Continuity of care interventions are defined as those focusing on the connection and coordination between patients and providers across time and settings and classified in informational, management, and relational continuity contexts (Reid et al., 2002; see Table 1).<br>**Comparator:** Usual care<br>**Outcome measures:** Hospital readmissions in time sections of | **Number of included studies & location:**<br>36 studies of which 30 in meta-analysis (3 in UK). Global and no language limits<br>**Search dates:** From inception to 27 Jan 2019<br>**Key characteristics:**<br>All RCTs. 8,920 patients – 53% with chronic heart failure, 10% chronic obstructive pulmonary disease, 7% COPD and heart failure, 3% chronic lung disease. No other details | Two critical weaknesses: No justification of excluded studies. Both high and moderate quality studies were included in the analysis without testing risk of bias (RoB)<sup>5</sup> on effect size in sensitivity analyses.<br>Non-critical weaknesses; the search strategy may not be highly sensitive; Heterogeneity noted and discussed but no satisfactory explanation as to why it increased with the study time frames. Critically Low Quality | At 1 month from discharge, the continuity interventions were associated with lower readmission rates in 207/1595 patients in the experimental group (12.9%), versus 264/1645 patients in the control group (16%) (Relative Risk [RR], 0.84 [95% CI, 0.71, 0.99] p=0.04). From 1 to 3 months, readmission rates were lower in 325/1480 patients in the experimental group (21.9%), versus 455/1523 patients in the control group (29.8%) (RR 0.74 [0.65, 0.84] p<0.00001). A subgroup analysis showed that this positive effect was stronger when the interventions addressed all of the continuity dimensions (patient provider relationship, transfer and use of information, management continuity). After 3 months (from 3-6 months) this impact became inconclusive with moderate/high statistical heterogeneity.<br>*Note:* Although not included by authors in the abstract, data for 6-12 months suggested benefit (RR 0.84 (0.74, 0.95) p=0.007) |

<sup>5</sup> Risk of bias is the preferred term used by Cochrane for quality appraisal and the key concept of the Cochrane risk of bias tool. It does not consider other concepts such as the quality of reporting, precision (the extent to which results are free of random errors), or external validity (directness, applicability or generalizability).

|  |  |  |  |  |
| --- | --- | --- | --- | --- |
|  | 1 month; >1-3 months, >3-6 months and 12+ months post discharge |  |  |  |
| **Gonçalves-Bradley et al. 2017**<br>Early discharge hospital at home. Cochrane Database of Systematic Reviews. 6: CD000356. doi: 10.1002/14651858.CD000356.p ub4 | **Review aim:** To determine the effectiveness and cost of managing patients with early discharge hospital at home compared with inpatient hospital care.<br>**Population:** Adults aged 18+ discharged from acute hospital inpatient care<br>**Intervention:** Early Discharge Hospital at Home (Descriptions vary: In the UK the focus is usually on the provision of personal, nurse-led care, building on the existing structure of primary care.<br>**Comparator:** Acute Hospital In-patient Care<br>**Outcome measures (older people):** Mortality, hospital readmissions, functional status, length of stay, living in institutional setting | **Number of included studies & location:**<br>Global but studies mainly conducted in high income countries [32 RCTs, 16 in UK].<br>Studies with data on older people aged 65+ = 16 RCTs (8 in the UK)<br>**Search dates:** From inception to 9 Jan 2017<br>**Key characteristics:**<br>RCTs with data for older people (65+):<br>Caplan 2006, Cotton 2000 [UK], Crotty 2002, Cunliffe 2004 [UK], Diaz Lobato 2005, Donald 1995 [UK], Harris 2005, Ince 2014, Martin 1994 [UK], Ojoo 2002 [UK], Rada 2008, Richards 1998 [UK], Shepperd 1998 [UK], Skwarska 2000 [UK], Tibaldi 2013, Utens 2012 | Critical domains:<br>Cannot exclude publication bias, no formal investigation of heterogeneity. No subgroup analysis to explore low and unclear risk of bias in studies and no discussion of this (but a vast review already). Claimed small likelihood of publication bias but didn't explore and no search for grey literature/unpublished studies.<br>Non-critical: No expert contacts or grey literature search; no assessment of sources of funding for trials.<br>Critically Low Quality | *Sections only included: Multiple relevant analyses are included for older people (older than 65) See pp.15-16 for narrative and pp.81 – 91; 100-101 for Forest plots.*<br>Authors concluded that the review provided low- to moderate-certainty evidence that hospital at home does not adversely affect mortality, hospital readmission, or functional status. The following conclusions were drawn based on the findings for people aged 65+:<br>*Mortality:*<br>Early discharge hospital at home probably makes little or no difference in mortality to older people with a mix of conditions, or COPD. Twelve trials reported data for mortality at three to six months follow-up for older people with a mix of conditions, and we pooled data from eight of them (RR 1.07 (95% CI 0.76, 1.49) p=0.71) and from five trials recruiting patients with COPD (RR 0.53 (0.25, 1.12) p=0.10).<br>*Hospital readmission:*<br>Early discharge hospital at home probably increases the risk of readmissions for older people with a with a mix of conditions, and may decrease the risk of readmissions for people with COPD. Pooling data for nine trials recruiting older people with a mix of conditions, median follow-up of three months (RR 1.25 (95% CI 0.98, 1.58; p=0.07), and five trials for participants with COPD with two to three months follow- up (RR 0.86 (0.66,1.13) p=0.29).<br>*Functional status:*<br>Based on the findings of four trials, Early discharge hospital at home probably makes little or no difference to functional status (Barthel Index) (mean difference 0.34 (95% CI -0.18, 0.86) p=0.20). There was substantial heterogeneity between trials.<br>*Patient reported outcomes:*<br>There was little or no difference between groups for older people with a mix of conditions and patients with COPD on a<br>range of patient reported outcomes (data provided in Analysis 2.7).<br>*Institutional care:*<br>Based on two trials with one year follow up and one with 6 months) the RR was (just significantly) in favour of early discharge re institutional care at one year: Risk Ratio 0.69 (95% CI 0.48, 0.99); p=0.04.<br>*Patient satisfaction:*<br>Of six trials reporting patient satisfaction, two reported increased levels of satisfaction for those allocated to early discharge hospital at home and four trials reported little or no difference.<br>*Caregiver outcomes:*<br>Five trials measured caregiver outcomes, including strain and general health. Three reported little or no difference (Cunliffe 2004; Shepperd 1998; Utens 2012), while two found less caregiver strain in early discharge hospital at home (Harris 2005; Tibaldi 2013). Three trials reported that early discharge hospital at home may increase carer satisfaction (Harris 2005; Ojoo 2002; Utens 2012), and two reported little or no difference (Caplan 2006; Shepperd 1998).<br>*Staff views:*<br>One trial reported that staff perceived that providing care in the patients' homes facilitated participation in rehabilitation, that the service was better staffed than the usual discharge services provided, and that rehabilitation services were coordinated with social care (Cunliffe 2004; results not tabulated); and a second trial reported little or no difference in general practitioners' level of satisfaction (Caplan 2006).<br>*Length of hospital stay:*<br>Combining data for four trials found that early discharge hospital at home probably reduces hospital length of stay (mean difference -6.76 days (95% CI -10.60, -2.92) p=0.0006. There was substantial heterogeneity. Pooling data from three trials that reported both length of stay in hospital and hospital at home suggested that early discharge hospital at home may increase the number of days of health care received (mean difference 6.43 (2.84,10.03) p=0.0005).<br>*Use of healthcare resources and cost:* |
|  |  |  |  | Seven trials reported the costs associated with the intervention, varied outcomes for different settings and populations suggesting that hospital at home may be more, less or the equivalent expense as in-hospital care. |
| **Gonçalves-Bradley et al. 2022**<br>Discharge planning from hospital. The Cochrane database of systematic reviews. 2: CD000313. doi: 10.1002/14651858.CD000313.p ub6 | **Review aim:** To assess the effectiveness of planning the discharge of individual patients moving from hospital.<br>**Population:** All patients in hospital (acute, rehabilitation or community) irrespective of age, gender or condition.<br>**Intervention:** Individualised discharge plan for a patient prior to them leaving hospital for home or residential care<br>**Comparator:** Routine discharge<br>**Outcome measures:** For older people – Mortality, hospital length of stay, readmissions | **Number of included studies & location:** global, 33 randomised trials (5 from UK).<br>Studies with data on older people = 16 (2 from UK)<br>**Search dates:** from inception to April 2021<br>**Key characteristics:**<br>Studies with data for older people:<br>Gillespie 2009, Goldman 2014, Harrison 2002, Lainscak 2013, Laramée 2003, Legrain 2011, Lindpaintner 2013, Moher 1992, Naughton 1994, Naylor 1994, Nazareth 2001 [UK], Nguyen 2018, Preen 2005, Rich 1993, Rich 1995, Sulch 2000 [UK] | There were a couple of non-critical weaknesses; no real discussion of heterogeneity (but trials grouped to minimise effects) and no account of risk of bias in the analysis (this was treated as non-critical since most of the included trials were assessed as low risk of bias).<br>Moderate Quality<br>Note: Older people is not defined but assumed 65+ as for Gonçalves-Bradley 2017. | *Sections only included:* three analyses are included for older people pp. 15 for text, pp.82, 86 for Forest plots.<br>A structured discharge plan that is tailored to the individual patient probably brings about a small reduction in the initial hospital length of stay and readmissions to hospital for older people with a medical condition and may slightly increase patient satisfaction with healthcare received.<br>*Mortality:*<br>For older people with a medical condition (usually heart failure), from pooling 8 trials, it is uncertain if discharge planning has an effect on mortality at three- to nine-month follow-up (RR 1.05 (95% CI 0.85, 1.29) p=0.67).<br>*Hospital length of stay*<br>An analysis of 11 trials of older people found a mean difference in hospital length of stay that just favoured discharge planning (mean difference -0.73 (95% CI -1.33, -0.12) p=0.02. For two trials looking a hospital length of stay following surgery there was no difference; mean difference = -0.06 (-1.23, 1.11) p = 0.92.<br>*Readmissions:*<br>For older people with a medical condition, discharge planning led to a relative reduction (from 17 trials) in readmissions to hospital (average follow-up within three months; risk ratio (RR) 0.89 (95% CI 0.81, 0.97) p=0.01). |
| **Handoll et al. 2021**<br>Multidisciplinary rehabilitation for older people with hip fractures. The Cochrane database of systematic reviews. 11: | **Review aim:** To assess the effects of multidisciplinary rehabilitation, in either inpatient or ambulatory care settings, for older people with hip fracture.<br>**Population:** Older people with any type of fracture of the proximal femur that had been surgically fixed prior to entry into the care programme. **Specific age limits not defined, but as anticipated, most participants** | **Number of included studies & location:** 28 trials from around the globe including four from the UK. Extracted studies: seven, with one from UK.<br>**Search dates:** 2009 (previous review) to Oct 2020 (trial registers only to Nov 2019).<br>**Key characteristics (of extracted section):** | This is an update of a review, with the protocol available on the Cochrane website. Deviations to the protocol identified.<br>Reason for choosing only randomised trials and trials using quasi-randomisation techniques not given. | **Extracted section findings:**<br>**Supported discharge and multidisciplinary home rehabilitation vs. usual care**<br>*People mainly living at home:* three trials (total N=377; Crotty et al 2003, Karlsson et al 2016; Ziden et al 2008). Very low-certainty evidence, so very little confidence in findings of little to no between-group difference in<br>poor outcome (death or move to a higher level of care or inability to walk) at three mo. (1 study) one year (3 studies);<br>quality of life at one year (1 study); |
| CD007125. doi: 10.1002/14651858.CD007125.p ub3 | **were aged 65 years and over.** Younger participants (<65 years) were included, as the number of these was relatively small. Trials of proximal femoral fracture in younger (<60) or those with multiple trauma were excluded, as were trials of people with metastatic disease or high-energy fractures. However, mixed population trials were included as long as participants in those excluded categories was relatively small with an unbiased distribution in trial arms or data were presented separately for the review target population.<br>**Intervention:** treatment in a multidisciplinary rehabilitation programme (where rehabilitation is delivered by a multidisciplinary team, supervised by a geriatrician, rehabilitation physician or other appropriate physician (or strong component of physician involvement), as opposed to 'usual' care (control group)). Such a programme would aim to improve the functioning of the person with hip fracture. The programme needed to have been provided in an inpatient or ambulatory setting (or both). Here, ambulatory setting covers home (including nursing homes), outpatient department and day hospital locations.<br>**Comparator:** Usual care (usual orthopaedic or medical care, or | Seven trials where care was provided predominantly in the community (in the community ('community') or mixed hospital and community care ('mixed')):<br>*Supported discharge and multidisciplinary home rehabilitation vs. usual care*<br>• Crotty 2003 (mixed) n=66<br>• Karlsson 2016 (mixed) n=209<br>• Ziden 2008 (mixed) n=102<br>• Crotty 2019 (community) n=240<br>*Intensive vs. less intensive multidisciplinary home rehabilitation:*<br>• Ryan 2006 (community; UK) n=71<br>*Multidisciplinary care, including progressive resistance training for one year vs. usual care*<br>• Singh 2012 (community) n=124<br>*Post-discharge multi-disciplinary clinic, with referral for further rehabilitation as necessary vs. usual care*<br>• Ashe 2019 (community) n=53<br>All seven were randomised trials (including one RCT). Sample sizes ranged from 53 to 240. | Search strategy comprehensive, but no details of whether content experts consulted.<br>Study selection (except for initial screen) and data extraction done by at least two reviewers.<br>Included studies described in detail, including funding sources, and excluded studies listed with reasons.<br>RoB assessed using tool outlined in the Cochrane Handbook for Systematic Reviews of Interventions.<br>Appropriate meta-analysis methods used and potential impact of RoB on analyses explored for the primary outcome ('poor outcome') in the main analysis using sensitivity analysis.<br>Impact of RoB included in the discussion; GRADE used and level of certainty downgraded due to RoB.<br>Heterogeneity (or lack of) in the results discussed.<br>Publication bias assessed. | • mortality at 4 or 12 months (2 & 3 studies respectively);<br>• independence in personal activities of daily living (1 study);<br>• moving permanently to a higher level of care by 12 mo. (2 studies); or<br>• being unable to walk at 3 or 12 months (1 & 2 studies respectively).<br>Long-term hip-related pain was not reported in any trial.<br>Further data was presented about assessment of dependency. Statistically significant findings of a positive intervention effect from one study for the modified Barthel Index at 4 mo. (analysis not presented and clinical importance not established). All very low-certainty evidence.<br>Evidence for all the other outcomes is of very low certainty.<br>• No difference in medical complications or delirium after discharge (1 study)<br>• No difference in number readmitted with 4 or 12 mo. (2 studies).<br>• Very low certainty evidence of little to no difference in fall and further fracture at 4 (1 study) or 12 mo. (1 study).<br>• Care burden at 4 mo. had a median difference of one-point, and at 12 mo. a three-point difference (p=0.02) favouring home rehabilitation (1 study).<br>• Shorter hospital stay for home-based rehabilitation participants by 6 days (2 studies) and mean length of stay over a day shorter but not statistically significant (1 study). (authors note duration of rehabilitation would have been longer in intervention groups).<br>• Home rehabilitation group spent 376 fewer days in institutional care than usual care at 1 mo. (1 study).<br>• No difference in carer time, GP visits or use of community services at 4 mo. (1 study).<br>*Nursing home residents:* One trial (n=240; Crotty et al 2019). Low-certainty evidence that there may be no or minimal between group differences in<br>• 'poor outcome' defined as dead or unable to walk at 12 months; or |
|  | potentially a rehabilitation programme of lesser intensity, or with different components, to the intervention under study).<br>**Outcome measures:**<br>*Critical/main outcomes:* 'poor outcome' between 4 and 12 mo. (death or deterioration in functional status leading to increased dependency in the community or admission to institutional care), health-related quality of life, mortality (all-cause) at ~4 & ~12 mo., dependency in activities of daily living at ~4 & ~12 mo., mobility between 4 & 12 mo., pain ~4 & ~12 mo..<br>*Other important clinical outcomes:*<br>'poor outcome' at hospital discharge, residential status between 4 & 12 mo., medical complications/sequelae, readmission, carer burden and stress. For ambulatory settings additional outcomes: number of fallers, number of fall-related hospital admissions and new fractures.<br>*Economic outcomes:* costs and resource data, length of hospital stay, overall length of secondary care stay, subsequent admission rates to, and days spent in, institutional care. |  | Conflict of interest and funding declared.<br>Moderate quality. | • in mortality at 4 months or 12 months (but very low certainty of fewer deaths at 4 weeks in the intervention group).<br>Due to very low-certainty evidence, we have very little confidence in the findings of no between-group differences in<br>• dependency at 4 weeks or at 12 months (slightly higher at one month for intervention group but not known if clinical important); or<br>• quality of life (with differing direction of effects with different measures), inability to walk or pain at 12 months.<br>Very low-certainty evidence for more falls in the intervention group at 4 weeks.<br>Mean per participant 12-month Australian Medicare costs at 2015/16 unit prices were not statistically significant. Analyses by the trial authors were in favour of the intervention group, but they noted that the Australian regulatory bodies would not deem it cost-effective due to their thresholds.<br>**Intensive vs less intensive multidisciplinary home rehabilitation (1 study, Ryan et al 2006):**<br>Evidence for all reported outcomes for those with hip fracture is of very low certainty. No evidence of differences between the two groups on any measure ('poor outcome', mortality, institutional care, QoL, dependence and activities).<br>**Multidisciplinary care including progressive resistance training for one year, vs. usual care (1 study, Singh et al 2012)**<br>Results were very low certainty evidence. Fewer participants had a 'poor outcome' in the intervention group at 12 mo. Dependency was marginally better in the intervention group at 12 mo. and assistive device use was lower in the intervention group. There was no difference in assessment of living skills and resources.<br>**Post-discharge multidisciplinary clinic with referral for further rehabilitation as necessary vs. usual care (1 study, Ashe et al 2019)** |
|  |  |  |  | Only very limited outcome data (physical activity and sedentary behaviour) from this study, none of which featured within the review.<br>**Notes:** authors note that the characteristics of multidisciplinary rehabilitation and usual care varied considerably. Results need to be interpreted in light of the clinical heterogeneity in intervention, populations and measures.<br>The seven trials in the extracted section all had lower age inclusion criteria of at least 65 years except Singh et al. 2012 which had a lower age limit of 55 years (mean age of sample was 79.3 years). |
| **Langhorne et al. 2017**<br><u>Early supported discharge services for people with acute stroke.</u><br>The Cochrane database of systematic reviews. 7: CD000443. doi: 10.1002/14651858.CD000443.p ub4 | **Review aim:** To establish if, in comparison with conventional care, services that offer people in hospital with stroke a policy of early discharge with rehabilitation provided in the community (ESD) can: 1) accelerate return home, 2) provide equivalent or better patient and carer outcomes, 3) be acceptable satisfactory to patients and carers, and 4) have justifiable resource implications use.<br>**Population:** Stroke patients in hospital (average ages ranged from 60-80).<br>**Intervention:** Any service intervention that has provided rehabilitation and support in a community setting with an aim of reducing the duration of hospital care<br>**Comparator:** usual/conventional care (control services were categorised on whether organised stroke unit | **Number of included studies & location:**<br>17 randomised trials (also three studies awaiting classification and seven ongoing studies) from around the globe, including four from the UK. Extracted studies: nine trials, four of which were from the UK.<br>**Search dates:** up until Dec 2016, Jan 2017 or Mar 2017.<br>**Key characteristics (of extracted section):**<br>Nine trials were included in the subgroup analysis which split the samples by age (<75 years and > 75 years) and examined the primary outcome (death or dependency; nine trials) and length of stay (eight trials <75 years and seven trials >75 years).<br>Sample sizes ranged from 23 to 331.<br>Multiple references were listed for studies, here is the major publications for each, as identified by review authors.<br>Anderson et al 2000 | No statement of review protocol, but this was an update. Changes in review listed in "difference between protocol and review" section.<br>Reason for choosing only randomised trials not given.<br>Comprehensive search strategy.<br>Study selection (at full text) and data extraction by two reviewers (where appropriate from publications; individual level patient data obtained from trialists).<br>Included studies described in detail (but funding details not provided) and excluded studies listed with reasons.<br>RoB assessed using Cochrane tool | **Extracted section:** Subgroup data for the primary outcome (death or dependency) were available for at least nine trials. Smaller amounts of data were available for death, death or institutionalisation, and length of stay. There was no significant association of participant age or gender with the apparent effect of the ESD service.<br>**Main results from abstract (for reference):** The ESD group showed reductions in the length of hospital stay equivalent to approximately six days (mean difference (MD) -5.5; 95% CI -3 to -8 days; $p < 0.0001$; moderate-grade evidence). The primary outcome was available for 16 trials (2359 participants). Overall, the OR for the outcome of death or dependency at the end of scheduled follow-up (median 6 months; range 3 to 12) was OR 0.80 (95% CI 0.67 to 0.95, $p = 0.01$, moderate-grade evidence) which equates to five fewer adverse outcomes per 100 patients receiving ESD.<br>**Notes:** Authors note that evidence quality was generally good, but many of the trial were from over ten years ago. However, changes to stroke services and therapies will only have affected a small proportion of patients.<br>They also note that when including studies with lower risk of bias this strengthened the results.<br>Broad inclusion criteria for interventions. |
|  | care was available to patients prior to discharge).<br>**Outcome measures:** primary: composite of end-point of death or long-term dependency recorded at the end of scheduled follow-up.<br>Death, physical dependency, place of residence, length of index hospital stay, readmission to hospital, total cost of service interventions.<br>Secondary outcomes: Activities of daily living score & extended score, subjective health status, mood, carer outcomes (mood and subjective health status), patient and carer satisfaction and/or service preference. | Ronning & Guldvog 1998<br>Donnelly et al 2004 - UK<br>Rudd et al 1997 - UK<br>Dey et al unpublished - UK<br>Mayo et al 2000<br>Rodgers et al 1997 - UK<br>Bautz-Holter et al 2000<br>Widen et al 1998 | Appropriate meta-analysis method and heterogeneity assessed (planned) but limited discussion of the latter.<br>Sensitivity analyses to investigate impact of RoB, but no further discussion of this.<br>Publication bias not assessed.<br>Declaration of interests for the two named authors; discussion of potential conflict (trialists who participated were also authors of the included trials).<br>Critically low quality. |  |
| **Lee et al. 2022**<br><u>Transitional care from hospital to home for frail older adults: A systematic review and meta-analysis.</u><br>Geriatric Nursing. 43: 64-76. doi: 10.1016/j.gerinurse.2021.11.003 | **Review aim:** To systematically review and synthesis the existing evidence on the effect of transitional care on all health outcomes of frail older adults who were discharged home from the hospital.<br>**Population:** frail older adults ≥ 65 years who were in transition from hospital to home.<br>**Intervention:** healthcare services that provided continuity of care. Focused on hospital-based transitional care, which was initiated during admission and continued after discharge. Intervention varied widely. Hospital based included geriatric assessment, discharge planning | **Number of included studies & location:**<br>21 studies reporting 14 trials from around the globe, including two from the UK.<br>**Search dates:** inception to August 2021.<br>**Key characteristics:**<br>Only RCTs included.<br>Total number of participants was 5776 with an age range of 77.0 to 85.7 years.<br>Mean sample size was 413 (range 128 to 2353).<br>Publication years were 1995 to 2021.<br>Berglund (2013)<br>Berglund (2015) | Reason for choosing only RCTs not given.<br>Registered on PROSPERO.<br>Search strategy described but more details needed (e.g. justification for restrictions, searching reference lists).<br>Two authors independently completed data extraction.<br>PRISMA provided with Ns of excluded studies with reason, but individual studies not listed. | **From abstract:** The meta-analysis demonstrated that transitional care reduced readmission at six months but not other time points nor mortality or quality of life. The intervention effectiveness was inconclusive.<br>**Results:** Results for the impact on readmission were mixed, with some studies reporting no effect (n=7) and others reporting lower readmission rates in the intervention groups (at varying timepoints; n=6). Pooled results showed a significant effect on readmission at six months (though not at other timepoints), but heterogeneity suggested ($I^2=81\%$).<br>There was no effect of interventions on mortality (n=9).<br>Findings were mixed for physical functioning; some trials showed improvements (n=3) but others reported no improvement (n=4).<br>Interventions had a significant impact on health-related QoL in two studies, but not in three others. Pooled results suggested no effect, but substantial heterogeneity was reported ($I^2=95\%$).<br>Four studies reporting on self-rated health and life satisfaction all showed interventions had a positive impact. |
|  | (more advanced or structured than usual), early rehabilitation and advanced care planning. Home based included home visits, rehabilitation, community care service, telephone follow-up, geriatric assessment and home safety assessment. Multidisciplinary approach and family involvement considered throughout.<br>**Comparator:** usual care provided by the hospital or clinic.<br>**Outcome measures:** all healthcare outcomes.<br>Included studies reported on the following: Readmission, quality of life, physical functioning, self-rated health, cost, life satisfaction, institutionalization, length of hospital stay, satisfaction to care, bed days, self determination, fall, co-morbidity, days at home. | Eklund (2013)<br>Ekelund (2015)<br>Ebrahimi (2017)<br>Wihelmsen (2017)<br>Courtney (2009)<br>Del Sindaco (2007)<br>Puligano (2010)<br>Finlayson (2018)<br>Edman (2013)<br>Tanajewski (2015) - UK<br>Hansen (1995)<br>Jepma (2021)<br>Lembeck (2019)<br>Nikolaus (1999)<br>Sahota (2017) - UK<br>Schapira (2021)<br>Siu (1996)<br>Reuben (1995)<br>Rytter (2010) | Included studies described in detail (but funding of individual studies not described) and RoB assessed.<br>Justification given for meta-analysis and heterogeneity assessed but causes not explored.<br>Potential impact of RoB on analysis/ interpretation not included.<br>Conflict of interest and funding declared.<br>Critically low quality. | Mixed results for the impact on costs; three trials reported lower medical costs in the intervention group, one that the intervention was cost-effective, but another that the intervention was not cost-effective.<br>No difference in nursing home placement reported by one trial, but those in the intervention group spent fewer days in nursing homes. There was no difference in institutionalization rate in one trial.<br>Satisfaction lower in the intervention group in one study but not significantly different in another (but which did show higher intervention quality).<br>**Notes:** Interventions varied widely (setting, healthcare providers, key coordinators, components and duration) as did outcome measures and timepoints for measurement.<br>Authors comment that higher intensity interventions appeared more effective.<br>They conclude that the effectiveness of transitional care interventions for frail older adults remains unclear. |
| **Mabire et al. 2016**<br><u>Effectiveness of nursing discharge planning interventions on health-related outcomes in discharged elderly inpatients: a systematic review.</u> JBI | **Review aim:** To determine the best available evidence on the effectiveness of discharge planning interventions involving at least one nurse on health-related outcomes for elderly inpatients discharged home and to assess the relative impact of individual components of discharge planning interventions.<br>**Population:** Elderly inpatients aged 65 years or older, discharged from acute care and | **Number of included studies & location:**<br>Thirteen studies from around the globe (none from the UK).<br>**Search dates:** 2000 to 2015<br>**Key characteristics:**<br>Ten RCTs, two pilot studies and one pre-post study.<br>A total of 3964 participants were included.<br>All over 60 years with a median of 77 years+ in four studies. | A priori protocol published.<br>Reason for study designs included not given.<br>Adequate search strategy, but justification for restrictions not given.<br>Two authors independently completed study selection (based on | **From abstract:** Nurse discharge planning did not significantly reduce hospital readmission rate (OR=0.73, 95% CIs=0.53-1.01, p=0.06). The overall effect score for length of stay was significant (weighted mean difference = 0.29, p<0.01), suggesting that discharge planning increased the length of hospitalization. The effectiveness of discharge planning did not significantly impact QoL (mental OR=0.37, p=0.19 and physical OR=0.47, p=0.15).<br>Findings of this review suggest that nursing discharge planning for elderly inpatients discharged home increases length of stay, yet neither reduces readmission rates nor improves QoL.<br>**Results:** |
| Database of Systematic Reviews and Implementation Reports, 14, 217–260. – doi:10.11124/JB ISRIR-2016-003085. | post-acute care rehabilitation hospitals to home.<br>**Intervention:** The review focused on the six keys components of Naylor's Transitional Care Model: early geriatric assessment, discharge preparation, patient or caregiver's participation, continuity of care, day of discharge assessment and post-discharge follow-up.<br>**Comparator:** usual care.<br>**Outcome measures:** length of stay, functional ability, symptoms management, adverse outcomes, unmet needs after discharge, coping with disease, health-related quality of life (QoL), satisfaction with care, readmission rate and healthcare utilization. | Follow up was from 0 to 31 months.<br>Anttila et al 2000<br>Arbaje et al 2010<br>Askim et al 2004<br>Carrington et al 2013<br>Counsell et al 2000<br>Courtney et al 2009<br>Dedhia et al 2010<br>Lim et al 2004<br>Naylor et al 2004<br>Forster et al 2005<br>Koehler et al 2009<br>Kwok et al 2005<br>Yu et al 2015 | critical appraisal) and data extraction.<br>Excluded studies at critical appraisal stage listed with reasons (but not all those read at full text) and PRISMA diagram presented.<br>Included studies described in partial detail; more detail about the populations, comparators, setting and funding of individual studies needed.<br>Details of what data used from non RCTs not given and meta-analyses results not presented separately for RCTs.<br>RoB assessed using JBI critical appraisal checklists but could have been more detailed (re selection of reported results).<br>Impact of RoB not assessed or discussed.<br>Subgroup analyses and discussion to explore explanations for heterogeneity of results.<br>Conflict of interest and funding declared.<br>Critically low quality. | *Readmission (10 studies):* Pooled results showed nursing discharge planning did not reduce hospital readmission, though heterogeneity was identified ($I^2=70.8\%$) and a funnel plot suggested possible publication bias. Sensitivity analyses removing particular studies changed the conclusions.<br>Subgroup analyses investigated the sources of heterogeneity:<br>• Type of intervention – all components of transitional care and follow up after discharge: both showed heterogeneity and results suggested neither interventions decreased readmission.<br>• *Types of providers* – nurse, case manager, advanced nurse practitioner and community nurse: three showed heterogeneity and results suggested no effect of provider type on hospital readmission.<br>• *Time to readmission* – one month and six or more months: both showed heterogeneity and results suggested measurement time had no effect on readmission.<br>• *Age* – younger than 77 years and older than 77 years: only studies of older people showed heterogeneity and results suggested no impact of age on readmission.<br>• *Country* – USA (n=3), Australia (n=3), China (n=2), Canada (n=1) & Finland (n=1): Australian studies showed heterogeneity and the results suggested that in the USA nursing discharge planning positively impacted readmission.<br>*Length of stay (6 studies):* Pooled analysis showed nursing discharge significantly increased length of stay. There was no significant heterogeneity. Sensitivity analysis omitting studies did not change the conclusions.<br>*QoL (3 studies):* Pooled analysis suggested no effect of nursing discharge planning on mental or physical QoL. Heterogeneity was identified on both dimensions of QoL.<br>**Notes:** A great deal of variation in the interventions across studies (post-discharge follow up was the most frequently reported intervention).<br>Authors suggest the strength of recommendations was low or very low.<br>Usual care not always described.<br>Substantial heterogeneity in providers and measures. |
| **Meulenbroeks et al. 2021**<br><u>The value of caregiver inclusive practices in geriatric transitions of care: A systematic review.</u> Health Policy 125; 7<br>doi:10.1016/j.healthpol.2021.05.001 | **Review aim:** To assess whether transitional care programs that integrate caregivers provide better value care than routine care<br>**Population:** People with geriatric syndrome (aged 65+ and displaying moderate to severe geriatric traits as measured by validated tool).<br>**Intervention:** Caregiver inclusive transitional care from acute to community setting<br>**Comparator:** Routine care<br>**Outcome measures:** Population health, healthcare professional experience, costs. | **Number of included studies & location:** 23 studies in high income countries (2 in the UK)<br>**Search dates:** Inception to June 2019<br>**Key characteristics:** 14 RCT, 9 NRCT. 90% rated as high or critical risk of bias. 16,657 patients in all. Average age 77.8 | No published protocol, search strategy unlikely to be comprehensive, single researcher only doing critical appraisal and data extraction; no discussion of heterogeneity.<br>Critically low quality | Narrative analysis only. Authors found that consistently positive results occurred for patient and caregiver satisfaction. Cost tended to increase with caregiver inclusive practices. Most studies found no difference in population health outcomes. They concluded that there was insufficient evidence on healthcare professional experience and insufficient evidence to determine whether caregiver inclusive transitions of care provide *better value* care than routine care. Studies that rigorously implement and evaluate caregiver inclusive care models are urgently required to inform future policy. |
| **Rasmussen et al. 2021**<br><u>Impact of transitional care interventions on hospital readmissions in older medical patients: A systematic review.</u> BMJ open. 11(1): e040057. doi: 10.1136/bmjopen-2020-040057. | **Review aim:** to evaluate the impact of transitional care interventions with both pre-discharge and postdischarge elements on readmission among older medical patients.<br>**Population:** older medical patients discharged from a general medical ward or emergency department. Patients aged ≥65 years or mean study population ≥75 years.<br>**Intervention:** in the transitional phase between hospital and home which examined the impact of the intervention on readmission rates. The | **Number of included studies & location:** Eleven studies from around the globe, one of which was from the UK.<br>**Search dates:** Jan 2008 to Aug 2019<br>**Key characteristics:** Five RCTs, four non-randomised controlled trials and two pre-post cohort studies. Sample sizes ranged from 41 to 19157, with a total of around 24500. Mean age of participants was ~78 (range 74.9 to 83.6) and ~79 (range 75.2 to 84.5) in the intervention and control groups respectively. | Reason for study designs included not given.<br>Registered on PROSPERO.<br>Comprehensive search strategy, but justification for restrictions not given.<br>Two authors independently completed study selection and data extraction.<br>Excluded studies listed with reasons and PRISMA diagram presented. | **From abstract:** Twenty-two of the 29 outcome results measuring re-admission rates (within 7–182 days after discharge) showed a drop in readmission rates in the intervention groups compared with the control groups. The most significant impact was seen when interventions were of high intensity, lasted at least 1 month and targeted patients at risk.<br>Methodological quality of included studies was reported as generally poor (two strong, three moderate and six weak).<br>**Results:** 22 of 29 outcome results showed the intervention to have a positive effect on readmission, however only 5 of the results (out of 29) were statistically significant. Eleven studies reported positive impacts of the intervention on readmission, one study reported both positive and no impacts, and three reported no impact. |
|  | interventions had to include both predischARGE and postdischarge components. The most pronounced difference between the interventions was in the predischARGE phase, whereas elements in the bridging and postdischarge phase were somewhat similar.<br>**Comparator:** usual care defined as standard care and treatment.<br>**Outcome measures:** unplanned readmission to hospital.<br>Subgroup analyses conducted of study population, intervention intensity, length of support, country of origin and outcome assessment. | Assessments were made 7-182 days after discharge.<br>Buurman et al 2016<br>Chow et al 2014<br>Courtney et al 2009<br>Finlayson et al 2019<br>Koehler et al 2009<br>Lin et al 2015<br>Nielson et al 2018<br>Robinson et al 2015<br>Rottman-Sagebiel et al 2018<br>Sahota et al 2017 - UK<br>Voss et al 2011 | Included studies described in partial detail; more detail about the populations, follow up and funding of individual studies needed.<br>RoB assessed but could have been more detailed (re selection of reported results).<br>Impact of RoB discussed.<br>Subgroup analyses and discussion to explore explanations for heterogeneity of results.<br>Conflict of interest and funding declared.<br>Low quality. | Subgroup analyses were completed for study population, intervention intensity, length of support, country and outcome.<br>• Positive impacts were greater and more commonly found among at risk patients compared with unselected patients.<br>• More high intensity interventions reported a positive impact on readmission than low intensity interventions. High intensity interventions had larger impacts and were statistically significant.<br>• Support of a month or more was associated with positive and statistically significant impacts (shorter length support had less or no impact).<br>• Great impact of the interventions was reported in studies based in non-European countries and all that were statistically significant were non-European.<br>• The impact was largest within 30 days of discharge. However, most statistically significant results were for outcomes assessed between one and three months.<br>**Notes:**<br>Caution for interpreting the results in that there were only 11 included studies and using GRADE the certainty of the evidence is low.<br>Several studies had small sample sizes and these showed stronger impact.<br>Authors note some trends in the components of interventions that had a positive impact on readmission, but not formally assessed.<br>The authors report that intervention fidelity was not reported by several studies and several studies did not adjust for confounders.<br>Language restrictions (English and Scandinavian languages) may have limited the scope.<br>Researchers contacted by the authors reported difficulty or decided not to publish no or negative results, thus negative findings may be under represented.<br>Ranges of participant ages displayed in the table of studies suggest one study included people from age 60 years. |
| **Tomlinson et al. 2020**<br><u>Successful care transitions for older people: a systematic review and meta-analysis of the effects of interventions that support medication continuity.</u> | **Review aim:** evaluating interventions, aimed at supporting successful transitions of care for older patients through enhanced medication continuity, using a taxonomy of components<br>**Population:** participants with a mean age of 65 years or older, who were being prepared for hospital discharge or who had a recent discharge (intervention provided within 1 month of discharge or on first post-discharge primary care visit).<br>**Intervention:** activities relating to medication that supported continuity<br>**Comparator:** Usual care<br>**Outcome measures:** hospital readmission, safe use of medication, quality of life. | **Number of included studies & location:** Global, 2/24 from UK<br>**Search dates:** Jan 2003 to September 2019<br>**Key characteristics**<br>Of those relevant to our question:<br>Nine interventions were commenced during admission and continued post discharge, bridging the transition Five of these involved nurse-delivered interventions, sometimes acting as 'transition coaches', to facilitate the patient's role in self-care. Three were pharmacist-led and one was multidisciplinary.<br>A further six studies evaluated interventions that commenced post-discharge, of which five were delivered by pharmacists. One study involved automatic electronic transfer of patient information to the primary care provider.<br>Overall, intervention delivery ranged from a single time point to 12 months post-discharge. The most intensive activity period was between discharge and 3 months post-discharge. | Published protocol in advance<br>Robust search strategy but no grey literature/expert consultation used<br>No details of excluded studies provided (reasons for exclusion are included)<br>"Studies were highly heterogeneous, drawn from varying populations, care settings and included different combinations of components and delivery time points."<br>"Most of the included studies contained methodological flaws, which affected their risk of bias assessment."<br>RoB not accounted for within meta-analysis<br>Funding of included studies not discussed. Moderate quality | **Interventions commenced post discharge**<br>"None showed a statistically significant reduction in hospital readmission and all were considered to be high quality"<br>In the only included UK study relevant to our question (a post-discharge intervention, Holland et al, 2005), readmission rate was 30% higher in the intervention arm $p=0.009$). Visual analogue scale QoL scores also favoured control.<br>**Interventions commenced during admission and continued post discharge**<br>"Five of these studies demonstrated a statistically significant reduction in all-cause hospital readmissions. All five interventions included follow-up (telephone, home visit or both) and education, continuing until seven to 180 days post-discharge. Four of these studies were considered to be at the highest risk of bias."<br>**Overall study results**<br>"The meta-analysis [of 19 studies included], stratified by component, demonstrated that the activities associated with reduced hospital readmissions were self-management education or coaching (RR 0.81 [0.74, 0.89]), telephone follow-up (RR 0.84 [0.73, 0.97]) and medication reconciliation (RR 0.88 [0.81, 0.96])."<br>"Interventions that bridged the transition for up to 90 days were more likely to support successful transitions and reduce adverse outcomes. These interventions used on average more components than those focusing solely on hospital admission or post-discharge time periods (6.2 versus 3.6 versus 3.8 respectively), reflecting their higher intensity and longitudinal nature"<br>"It is difficult to attribute success to individual components within bundles and our meta-analysis illustrates a modest overall effect size." |
|  |  | **Interventions commenced during admission and included post discharge support (no UK studies)**<br>Buurman 2010<br>Casas 2006<br>Chan 2015<br>Coleman 2006<br>Gillespie 2009<br>Huang 2005<br>Koehler 2009<br>Lee 2015<br>Ravn-Nielsen 2018<br>**Interventions commenced post discharge (1 UK study – Holland et al 2005)**<br>Ahmad 2010<br>Char 2017,<br>Gurwitz 2014<br>Haag 2016<br>Holland 2005<br>Tuttle et al 2018 |  | Therefore, these results cannot demonstrate causality and we cannot draw firm conclusion” |
| **Williams et al. 2022**<br><u>Early supported discharge for older adults admitted to hospital with medical complaints: a systematic review and meta-analysis</u> | **Review aim:** to explore the totality of evidence for the use of early supported discharge in older adults hospitalised with medical complaints.<br>**Population:** Studies were included if > 50% of the study population were older adults (aged > 65 years) who were admitted to the acute care setting for medical reasons.<br>**Intervention:** interventions aimed to accelerate patient discharge from hospital once medically stable, and providing patients with the necessary input in the community at the same | **Number of included studies & location:** 5 (New Zealand=2, UK=1, Australia=1, Germany=1)<br>**Search dates:** 1997 to Jan 2021<br>**Key characteristics:**<br>All included studies are RCTs<br>Caplan et al 2005<br>Cunliffe et al 2004<br>Harris et al 2005<br>Nikolaus et al 1999<br>Parsons et al 2018<br>Follow up periods varied from 90 days to 12 months post intervention. | Published protocol in advance<br>Robust search strategy but no grey literature/expert consultation used<br>Excluded studies and reasons for exclusion provided in full<br>Risk of bias – all studies included rated as ‘some concerns’ – risk of bias not discussed elsewhere in report in relation to impact on results.<br>Funding of individual studies not discussed.<br>Critically low quality | “A significant reduction in length of stay associated with the implementation of early supported discharge among this population. (data from 4/5 included studies, \|Mean difference =-6.04, CI -9.76 to -2.32, p=0.001)<br>No statistically significant effects favouring ESD interventions were established in mortality (3/5 studies), function (3/5 studies), HRQoL (2/5 studies), hospital readmissions or long term care admissions or cognition (2/5 studies).”<br>Three studies reported on resource costs<sup>1</sup> found the intervention to be associated with higher costs, and two found the care of the control group was more costly.<br>Authors identify high levels of heterogeneity of the included studies relating to the interventions provided and the outcomes measured. |
|  | level of intensity and resources they would receive while in the inpatient setting. All interventions were MDT led, based in the participants homes, and included physiotherapists and OTs, with some also including a doctor, case manager or social worker. Intervention intensity and duration varied.<br>**Comparator:** Usual care<br>**Outcome measures:** length of hospital stay, functional abilities, quality of life, cognition, carer and patient satisfaction, unscheduled hospital readmissions (including frequency), nursing home admission, mortality and cost. | Average age of participants across all studies ranged from 79.8 to 83.8 (intervention), (controls – not detailed for every study age range 78.7 to 84). |  | Publication bias and/or meta-regression or subgroup analysis not conducted due to limited number of studies available for inclusion. |

**Abbreviations:** CI=confidence interval; COPD=Chronic obstructive pulmonary disease; ESD=early supported discharge; JBI=Joanna Briggs Institute; MDT= Multi-Disciplinary Team; mo.=months; NRCT= non-randomised controlled trial; OR=odds ratio; OTs=Occupational therapists; QoL=quality of life; RCT=randomised controlled trial; RoB=Risk of bias; RR=relative risk;

#### 6.2.2 Primary Research (Table 3)

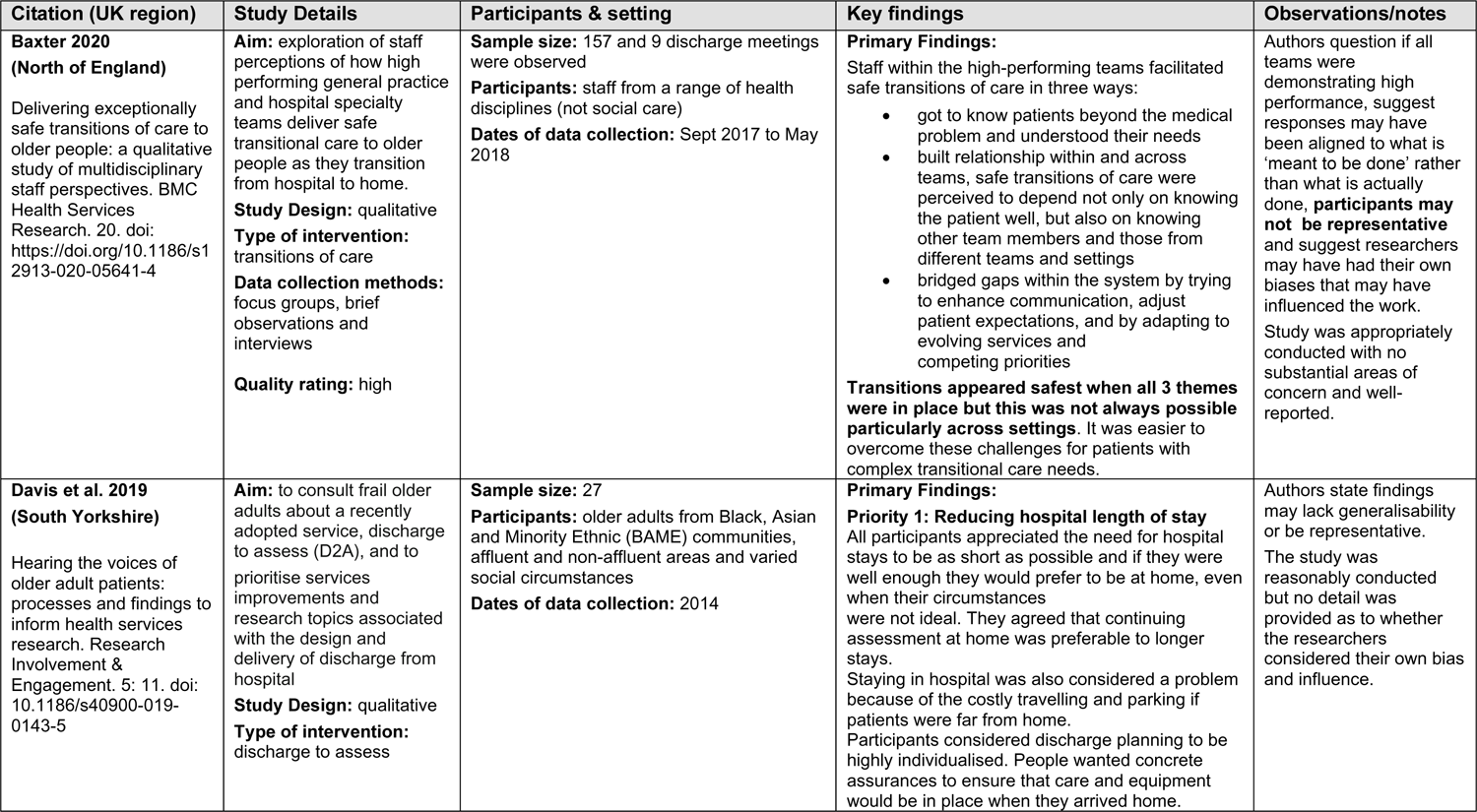
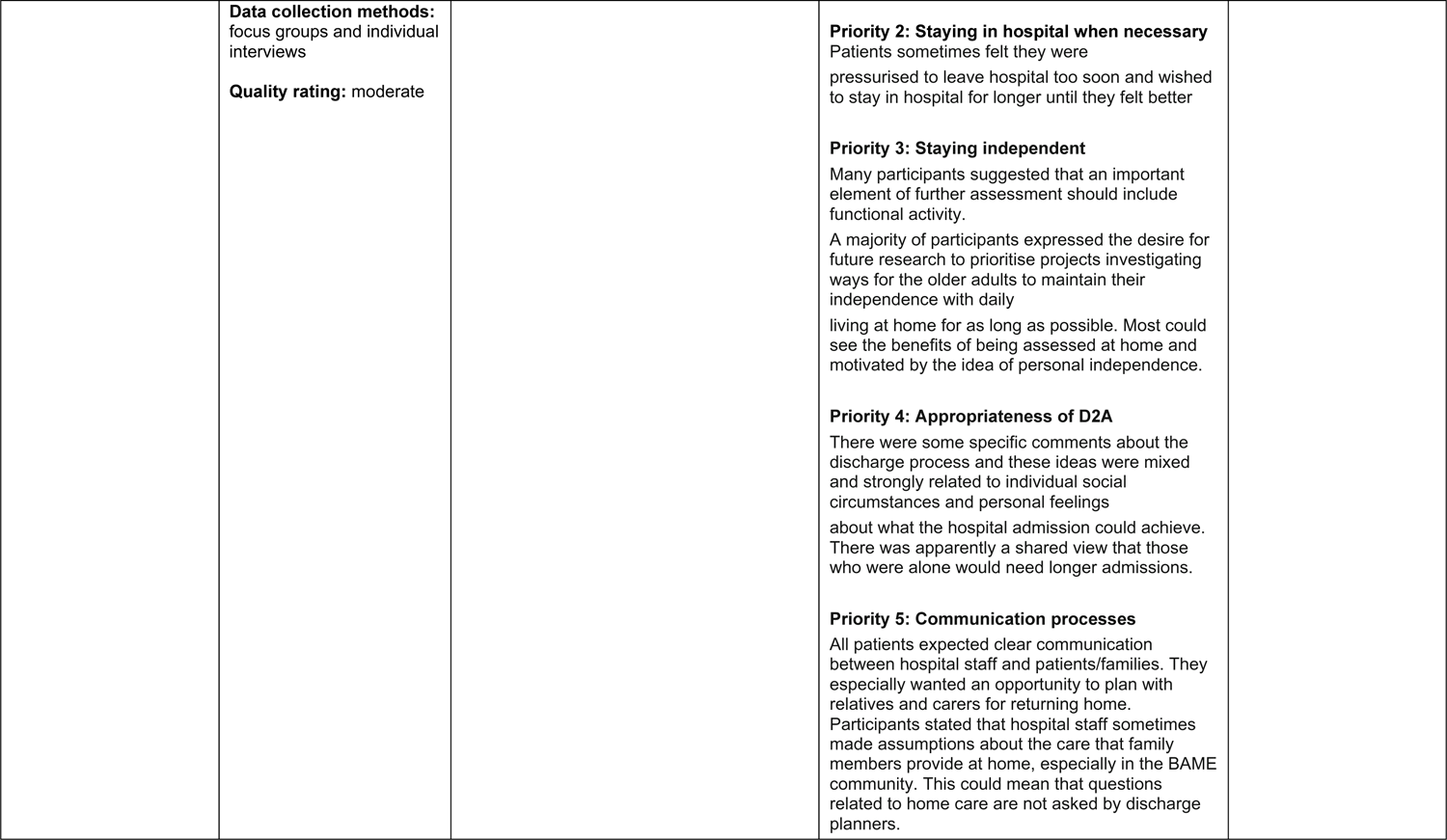
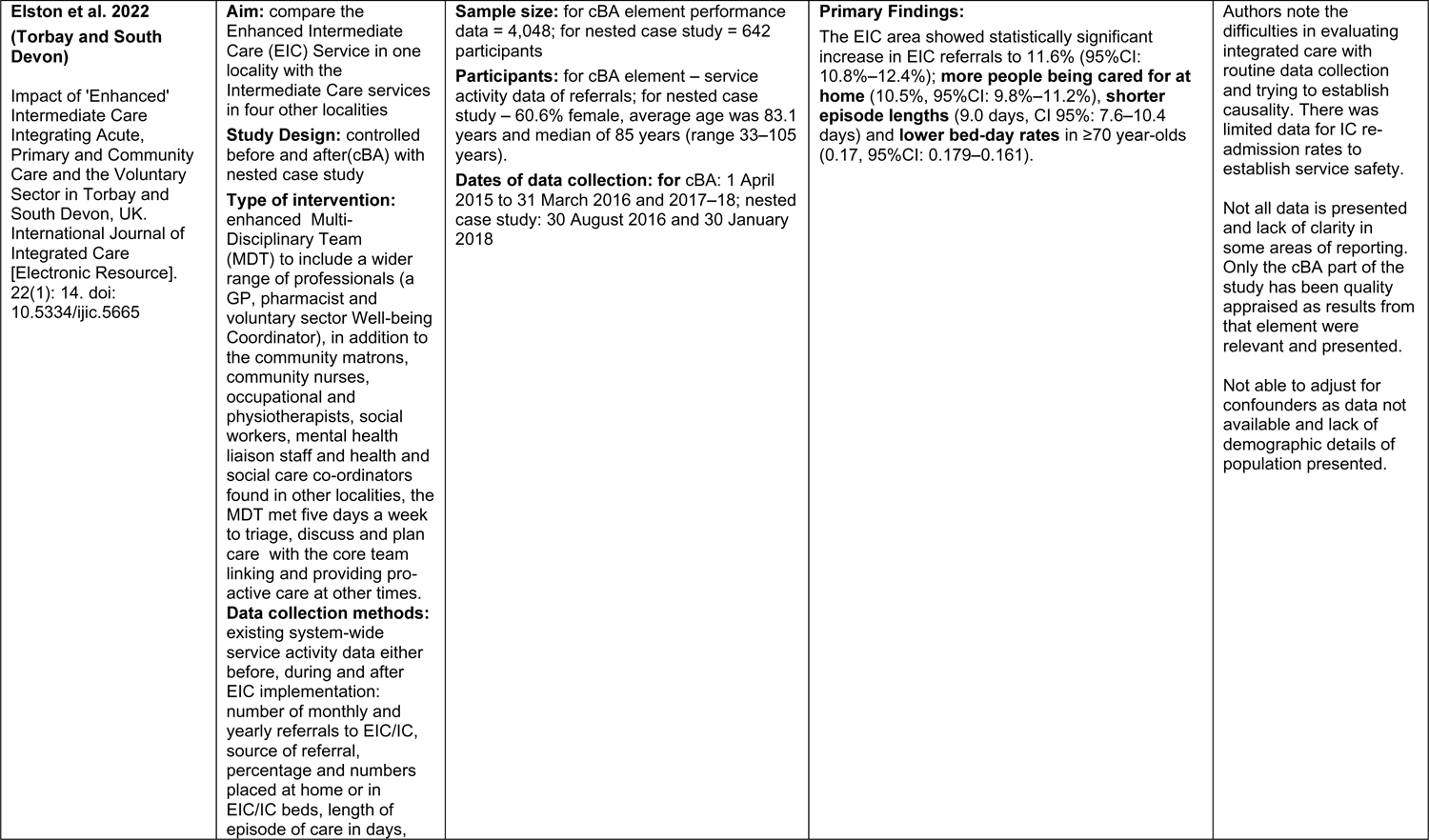
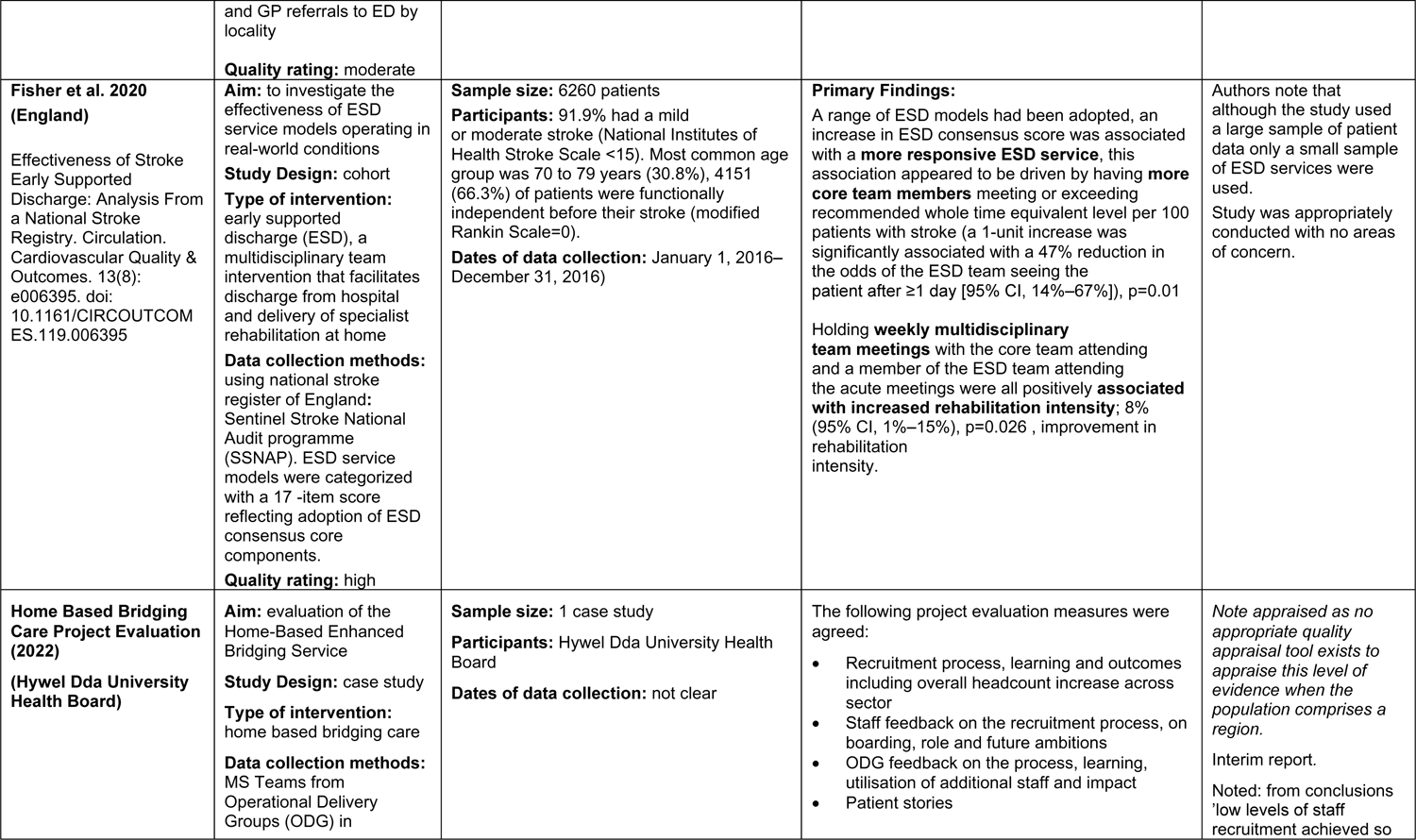
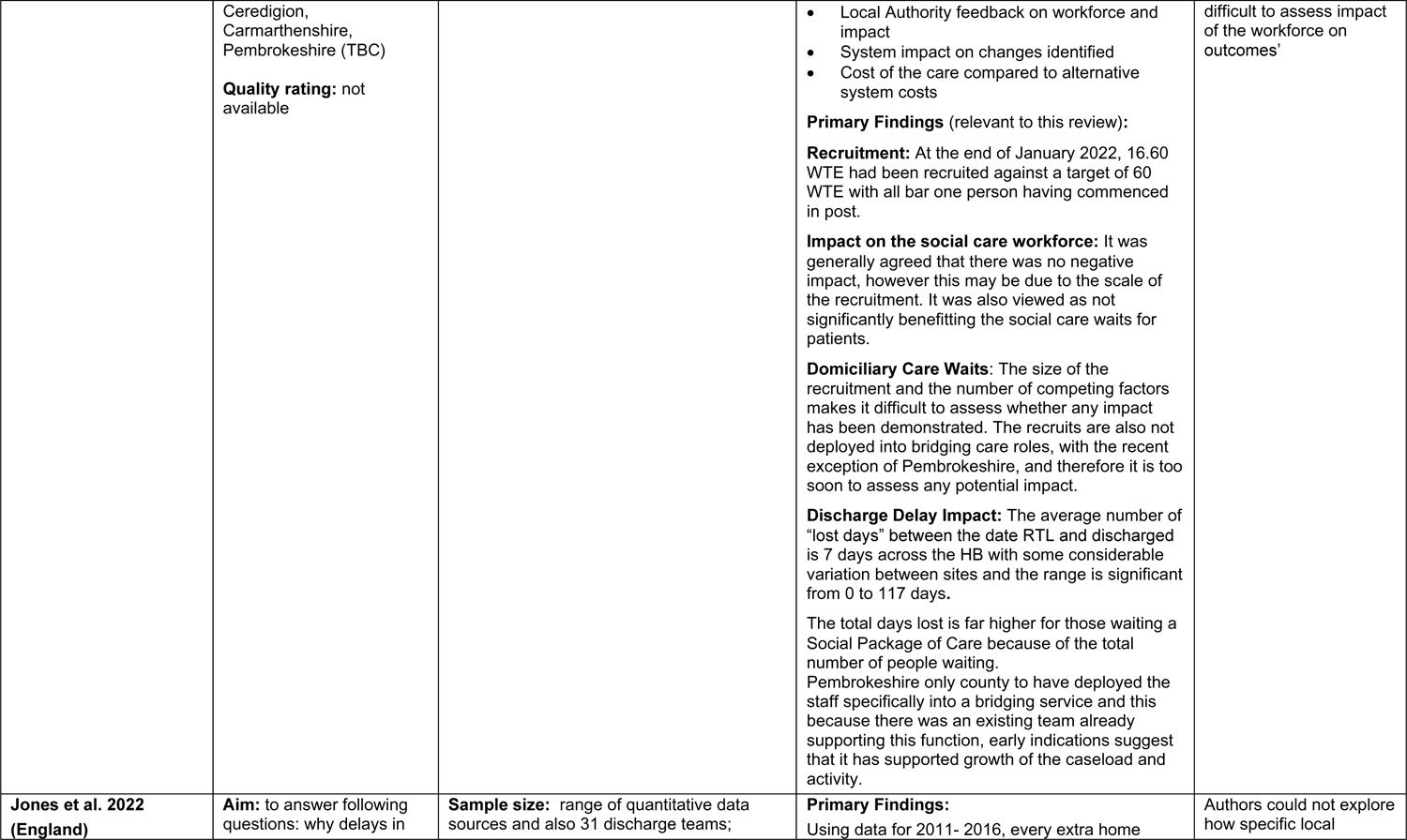
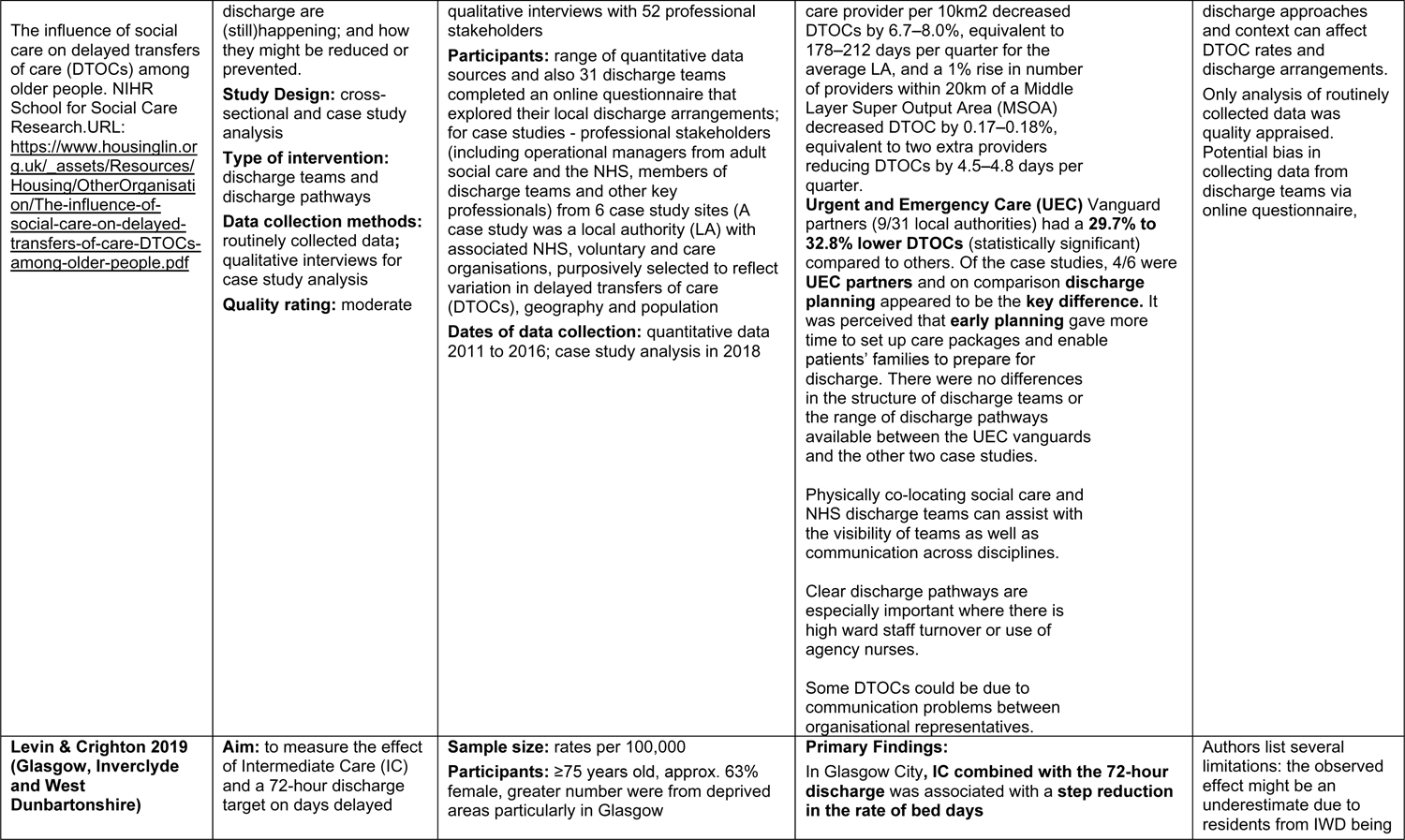
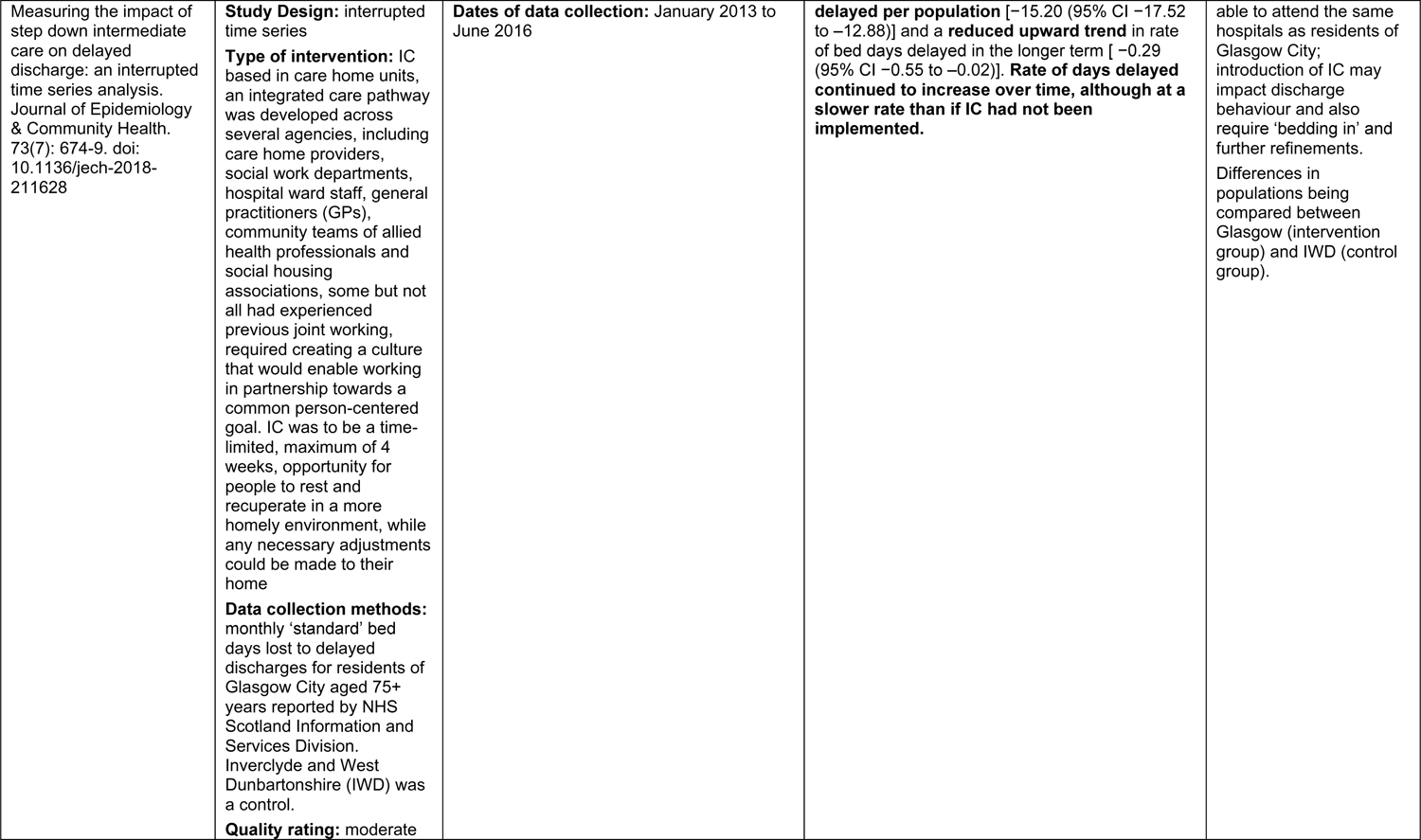
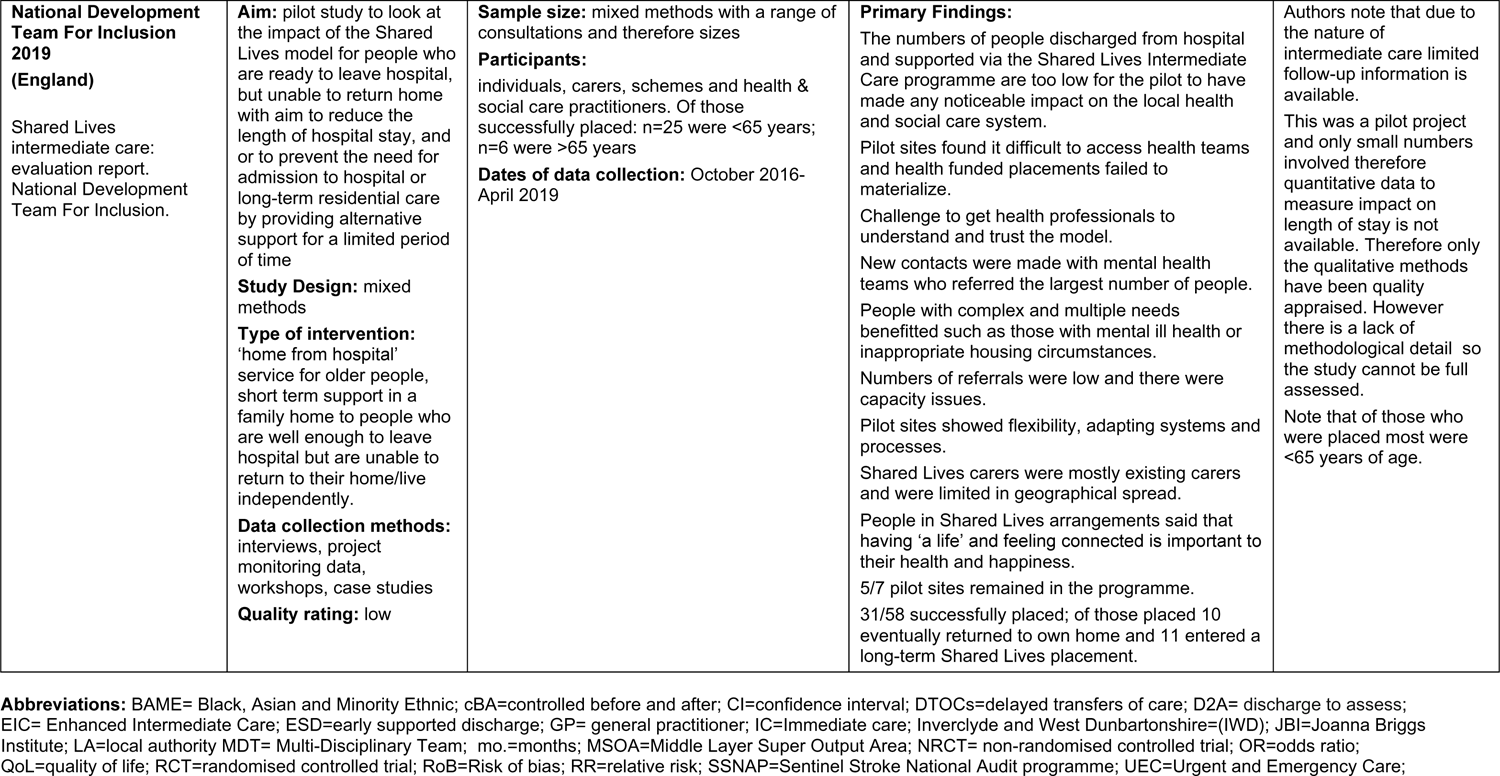

#### 6.2.3 UK primary studies of early supported discharge published since 2000 identified from systematic reviews (Table 4)

| Reference details | Readmission outcome | Hospital stays | Components |
| --- | --- | --- | --- |
| **Cotton** MM, Bucknall CE, Dagg KD, Johnson MK, MacGregor G, Stewart C, et al. <u>Early discharge for patients with exacerbations of chronic obstructive pulmonary disease: a randomised controlled trial</u>. Thorax 2000;55(11):902-6.<br>**RCT**<br>**Objective:** to compare early discharge with home treatment supported by respiratory nurses with conventional hospital management of patients admitted with exacerbations of COPD.<br>**Population:** patients with COPD admitted as emergencies (n=81); mean age circa 67 years. Glasgow.<br>**Bottom line:** Patients with acute exacerbations of COPD uncomplicated by acidotic respiratory failure or other medical problems can be **discharged home earlier than is current practice** with support by visiting respiratory nurses. No difference was found in the subsequent need for readmission. | **Twelve patients** (30% conventional management, 29.3% early discharge) were readmitted **in each group** giving a mean difference in readmission of 0.7% (95% CI of the difference -19.2 to 20.6) | **Early discharge reduced inpatient stay** from a mean of 6.1 (range 1–13) days with conventional management to 3.2 (1–16) days with an early discharge policy | Early discharge group was sent home on the next working day after recruitment (ideally within three days of admission)<br>**Patient discharge was not supported by increased use of social services support or rehabilitation services such as physiotherapy.** Pre-existing social services support was reinstituted if stopped before discharge.<br>Patients were visited by the respiratory nurse on the first morning after discharge and thereafter at intervals determined by the nurse.<br>Home management followed the practice developed for ARAS*<br>Both groups were recalled for review at the chest clinic two months after discharge (unless a current or recent inpatient).<br>*Flanigan U, Irwin A, Dagg K. An acute respiratory assessment service. *Professional Nurse* 1999;14:839–42. |
| **Cunliffe** A, Gladman JRF, Husbands SL, Miller P, Dewey ME, Harwood RH. <u>Sooner and healthier: a randomised controlled trial and interview study of an early discharge rehabilitation service for older people</u>. Age and Ageing 2004;33(3):246-52.<br>**RCT**<br>**Objective:** to evaluate an early discharge and rehabilitation service (EDRS) for older people.<br>**Population:** hospitalised older medical and surgical patients (n=370); mean age circa 79 years. Nottingham<br>**Bottom line:** some older people can be discharged from hospital sooner. | Number of patients re-admitted to hospital by 3 months<br>**EDRS – 49 (26%)**<br>**Control – 40 (22%)**<br>Relative Risk = 1.21 (0.85–1.76); no significant difference | Median, IQR (mean) length of stay from randomisation to discharge:<br>**EDRS - 6, IQR 4–13 (12)**<br>**Control - 13, IQR 6–24 (21)**<br>Median difference 4 (95% CI 3–7) | The EDRS was staffed by two occupational therapists, two physiotherapists, three nurses, a Community Care Officer (liaising with social services), seven rehabilitation assistants, and secretarial support. There were no doctors in the EDRS: medical care was given by the hospital team while in hospital and by the general practitioner when at home.<br>Up to four visits per day could be provided, up to 7 days per week, between the hours of 8am and 10pm. The package of care could last up to 4 weeks and was tailored to individual needs.<br>The EDRS provided a mean of 22 visits per participant. |
| **Donnelly** M, Power M, Russell M, Fullerton K. <u>Randomised controlled trial of an early discharge rehabilitation service: the Belfast Community Stroke Trial</u>. Stroke 2004;35(1):127-33.<br>**RCT**<br>**Objective:** to compare a community-based multidisciplinary stroke team (CST) approach with hospital-based rehabilitation.<br>**Population:** stroke patients (n= 113); mean age circa 70 years. Belfast<br>**Bottom line:** A mixed model of hospital-based and community-based rehabilitation services is likely to lead to increased patient choice and satisfaction and a potential reduction in bed pressures for less severe stroke patients. | There were a similar number of readmissions and the reason for readmission appeared to be related directly to stroke in 6 of 13 cases (*data not provided*) | There was an 8-day difference in mean inpatient stay (**P=0.304**) | The community-based multidisciplinary stroke team (CST) service consisted of a team comprising 0.33 coordinator, 1 occupational therapist, 1.5 physiotherapists, 1 speech and language therapist, and 2 rehabilitation assistants.<br>On average the number of home visits (each lasting 45 minutes) over a 3-month period was 2.5 per week.<br>Multidisciplinary meetings were held to discuss the assessment of patients and progress toward rehabilitation goals |
| **Ojoo** JC, Moon T, McGlone S, Martin K, Gardiner ED, Greenstone MA, et al. <u>Patients' and carers' preferences in two models of care for acute exacerbations of COPD: results of a randomised controlled trial</u>. Thorax 2002;57(2):167-9.<br>**RCT**<br>**Objective:** to identify patient and carer preferences for either hospital at home (HaH) or inpatient management<br>**Population:** patients with acute exacerbation of COPD (n=60); mean age circa 70 years. Hull<br>**Bottom line:** both patients and carers were significantly more likely to prefer domiciliary care if they were in the HaH arm | Readmission rate at 3 months (%)<br>HaH – 33.3<br>Inpatient care – 44.4<br>**No significant difference** | Mean no of days in care<br>HaH – 33.3<br>Inpatient care – 44.4<br>**No significant difference** | HaH – care from a team of two respiratory outreach nurses (RONs).<br>Patients were sent home within 48 hours of admission on a discharge package that included nebulised or inhaled bronchodilators, oral and inhaled steroids, antibiotics, and oxygen as necessary.<br>The patients' GPs were aware of, but were not involved with, the HaH patients. The RONS monitored the treatment of these patients daily and carried out patient and carer education and reassurance. |

**Abbreviations:** ARAS=acute respiratory assessment service; COPD=chronic obstructive pulmonary disease; CST=community-based multidisciplinary stroke team; EDRS=early discharge and rehabilitation service; HaH=hospital at home; RCT=randomised controlled trial; RONS=respiratory outreach nurses.

#### 6.2.4 UK primary studies of transitional care published since 2000 identified from systematic reviews (Table 5)

| Reference details | Readmission outcome | Hospital stays | Components (post discharge) |
| --- | --- | --- | --- |
| **Blue, L., Lang, E., McMurray, J.J., Davie, A.P., McDonagh, T.A., Murdoch, D.R., Petrie, M.C., Connolly, E., Norrie, J., Round, C.E., Ford, I., Morrison, C.E., 2001. Randomised controlled trial of specialist nurse intervention in heart failure. BMJ 323 (7315),715–718. doi: [10.1136/bmj.323.7315.715](https://doi.org/10.1136/bmj.323.7315.715)**<br>**RCT**<br>**Objective:** To determine whether specialist nurse intervention improves outcome in patients with chronic heart failure<br>**Population:** 165 patients with heart failure. Mean age circa 75. Glasgow.<br>**Bottom line:** Compared to usual (primary) care Specialist Nurse visits post discharge resulted in **significantly fewer re-admissions** for heart failure patients | Compared with usual care, patients in the **intervention group had fewer readmissions** for any reason (86 v 114, P = 0.018) [significant]<br>Taking the number of readmissions for each patient into account: The number of admissions/patient/month was 0.174 in the usual care group and 0.124 in the intervention group (**rate ratio 0.71, 95% confidence interval 0.54 to 0.94**) for all cause admissions; the corresponding rates for admission for heart failure were 0.069 and 0.027 (0.40, 0.23 to 0.71). [significant]<br>Time points: To 12 months | Patients in the intervention group spent fewer days in hospital for heart failure (mean 3.43 v 7.46 days, P = 0.0051). | Specialist Nurse: A number of planned home visits by nurse of decreasing frequency, to 12 months, supplemented by telephone contact as needed.<br>Educate patients on treatment, self monitoring and management, liaison with other health workers as required, psychological support<br>Usual care: Primary care |
| **Cleland, J.G., Louis, A.A., Rigby, A.S., Janssens, U., Balk, A.H., Investigators, T.-H., 2005. <u>Noninvasive home telemonitoring for patients with heart failure at high risk of recurrent admission and death: the Trans-European Network-Home-CareManagement System (TEN-HMS) study</u>. J. Am. Coll. Cardiol. 45 (10), 1654–1664. doi: 10.1016/j.jacc.2005.01.050.**<br>**RCT** | Number of hospitalisations (**differences between groups not analysed**)<br>240 days: HTM 155, NTS 144, UC 69<br>450 days: HTM 159, NTS 151, UC 85<br>Time points: To 240 & 450 days | Days in hospital for any reason over 240 days:<br>HTM vs NTS: -4 (-10, +2)<br>HTM vs UC: -1 (-9, +6)<br>NTS vs UC: = +5 (-2, +13)<br>[not significant]<br>Days in hospital for heart failure over 240 days:<br>HTM vs NTS: -1 (-5, +4)<br>HTM vs UC: 0 (-5, +5)<br>NTS vs UC: = -1 (-6, +5)<br>[not significant] | Home Telemonitoring (HTM): Twice-daily patient self-measurement of weight, blood pressure, heart rate, and rhythm with automated devices linked to a cardiology centre<br>Nurse Telephone support (NTS): Specialist nurses who were available to patients by telephone<br>Usual care (UC): Primary care |
| **Objective:** To identify whether home telemonitoring (HTM) (after discharge) improves outcomes compared with nurse telephone support (NTS) and usual care (UC) for patients with heart failure who are at high risk of hospitalization or death<br>**Population:** 426 patients with heart failure. Mean age 67-68. From UK, Germany, Netherlands<br>**Bottom line:** Compared to usual (primary) care home telemonitoring or nurse telephone support have no significant effect on days spent in hospital after discharge for heart failure patients |  | At 450 days, no significant differences |  |
| **Edmans J, Bradshaw L, Franklin M, Gladman J, Conroy S.** Specialist geriatric medical assessment for patients discharged from hospital acute assessment units: randomised controlled trial. BMJ. 2013;347:f5874. https://doi.org/10.1136/bmj.f5874<br>**RCT**<br>**Objective:** To evaluate the effect of specialist geriatric medical management on the outcomes of at risk older people discharged from acute medical assessment units<br>**Population:** 430 patients aged 70+ discharged within 72 of attending an acute medical assessment unit and at risk of decline. Nottingham and Leicester | Mean **hospital presentations for intervention versus usual care = 1.32 (1.01, 1.74) p = 0.05** [just significant]<br>Means **days at home for intervention versus usual care = -0.5 (-4.6, +3.6); p=0.31** [not significant]<br>Time points: 90 days |  | Specialist geriatric medical management: Agreed by geriatricians pre-discharge and seemed to comprise home visits, clinic visits, phone call, other patient related activity. Health care professional(s) not specified.<br>Usual care: Primary care |
| **Bottom line:** Compared to usual (primary) care, specialist geriatric medical management (unclear components) does **not affect overall days spent at home** but may lead to **a small increase in hospital presentations** for frail patients aged 70+<br>Linked paper:<br>**Tanajewski L, Franklin M, Gkountouras G, et al.** Cost-effectiveness of a specialist geriatric medical intervention for frail older people discharged from acute medical units: economic evaluation in a two-centre randomised controlled trial (AMIGOS). PLoS One. 2015;10:(5) e0121340. https://doi.org/10.1371/journal.pone.0121340. |  |  |  |
| **Holland R, Lenaghan E, Harvey I et al.** <u>Does home based medication review keep older people out of hospital? The HOMER randomised controlled trial.</u> Bmj. 2005;330(7486):293-0. doi: 10.1136/bmj.38338.674583.AE.<br>**RCT**<br>**Objective:** To determine whether home based medication review by pharmacists affects hospital readmission rates among older people<br>**Population:** 872 patients aged 80+ discharged after emergency admission for any cause. Norfolk and Suffolk<br>**Bottom line:** Compared to usual care (components unstated) two home visits by a pharmacist was associated with **a significantly** | 178 readmissions occurred in the control group and 234 in the intervention group (**rate ratio = 1.30, 95% confidence interval 1.07 to 1.58; P = 0.009**) [significant]<br>Time points: 6 months |  | Home based medication review: Two home visits by a pharmacist within 2 weeks and eight weeks of discharge to educate patients and carers about drugs, remove out of date drugs, inform GPs of drug reactions/interactions and inform local pharmacist if compliance aid needed.<br>Usual care: Not stated. Presume primary care |
| **higher rate of hospital re-admissions** for patients aged 80+ discharged after an emergency admission |  |  |  |
| **Sahota** O, Pulikottil-Jacob R, Marshall F, et al. The Community In-reach Rehabilitation and Care Transition (CIRACT) clinical and cost-effectiveness randomisation controlled trial in older people admitted to hospital as an acute medical emergency. Age Ageing. 2016;46(1):26–32. https://doi.org/10.1093/ageing/afw149.<br>**RCT**<br>**Objective:** To compare the clinical and cost-effectiveness of a Community In-reach Rehabilitation and Care Transition (CIRACT) service with the traditional hospital-based rehabilitation (THB-Rehab) service<br>**Population:** 250 frail older people aged 70+ admitted to hospital as an acute emergency. Nottingham<br>**Bottom line:** Compared to usual care (community services post-discharge) a specialist occupational therapy service with links to a social services practitioner does **not results in significantly reduced hospital lengths of stay or re-admissions** for frail older people following admission as an acute emergency | Of the participants who were discharged from hospital, 17% and 13% were readmitted within 28 days from the CIRACT and THB-Rehab services, respectively (**risk difference 3.8%, 95% CI –5.8% to 13.4%**) [not significant]<br>Time points: 28 days | There was **no significant difference** in length of stay between the CIRACT and THB-Rehab service (median 8 versus 9 days; geometric mean 7.8 versus 8.7 days, mean ratio 0.90, 95% confidence interval (CI) 0.74–1.10) [not significant but trend to a reduction with the CIRACT service] | Community In-reach Rehabilitation and Care Transition (CIRACT): Senior occupational therapist and assistant practitioner, linked directly to a social services practitioner making contact pre-discharge and continuing post-discharge care. Home visits to assess rehabilitation needs, follow up visits and referral to community services as required.<br>Usual care THB-Rehab: Community services (presumed primary care) post-discharge. [Pre-discharge = THB-Rehab ward based team of occupational therapist/physiotherapists] |

**Abbreviations:** CIRACT=Community In-reach Rehabilitation and Care Transition; HTM = home telemonitoring; NTS=nurse telephone support; THB-Rehab)traditional hospital-based rehabilitation; UC=usual care.

### 6.3 Quality appraisal of studies

#### 6.3.1 Secondary research (Table 6)

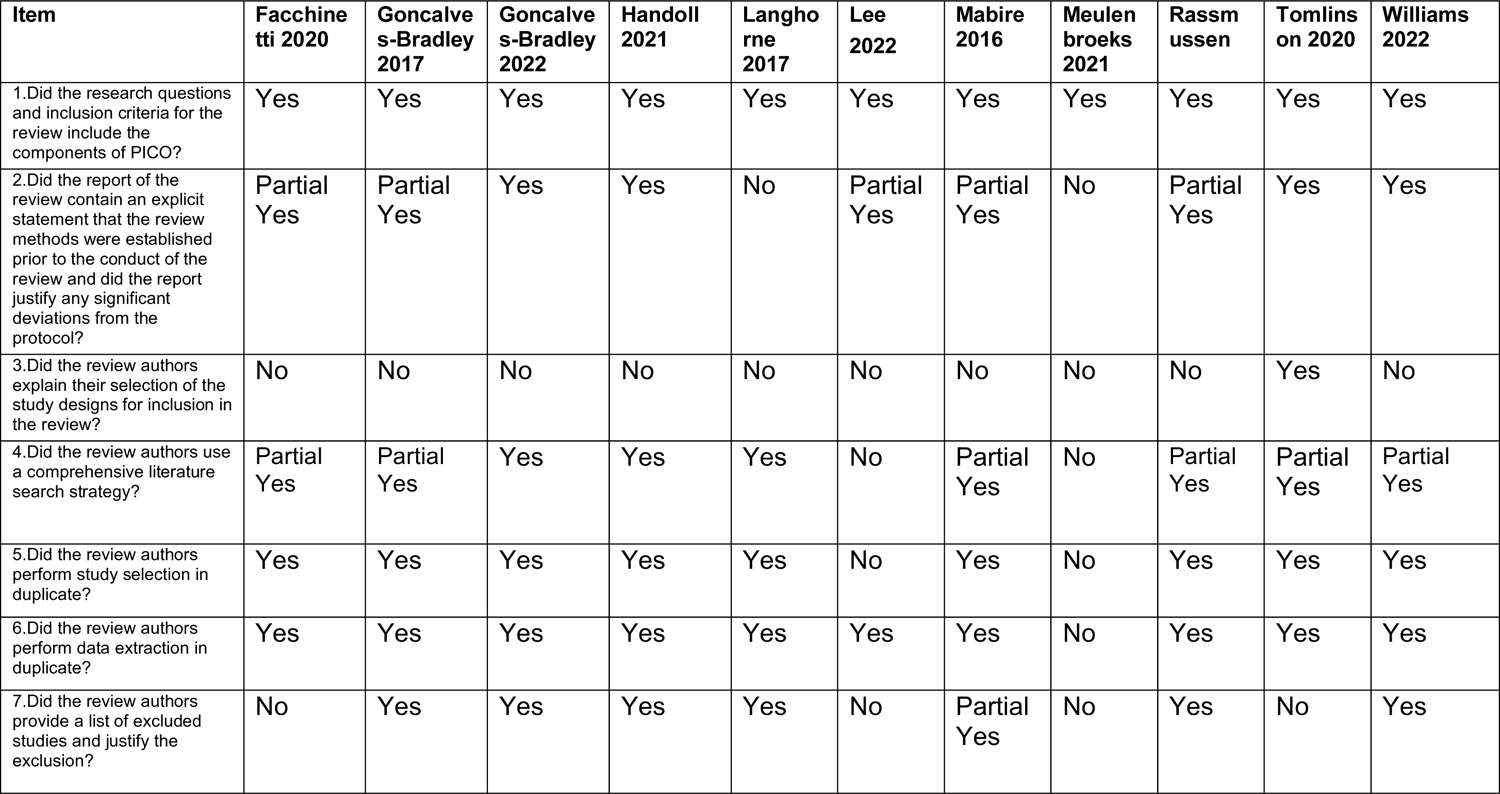
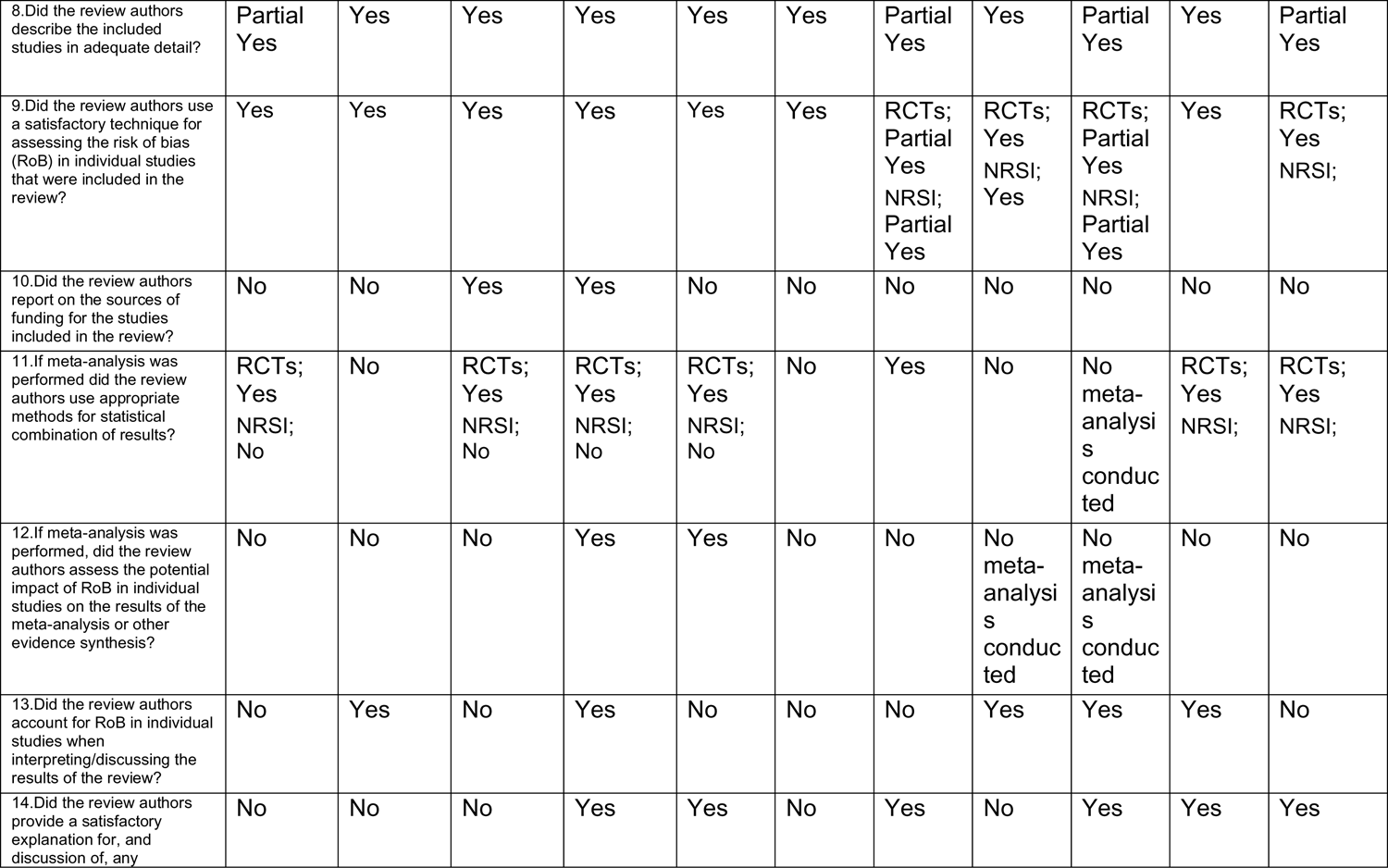
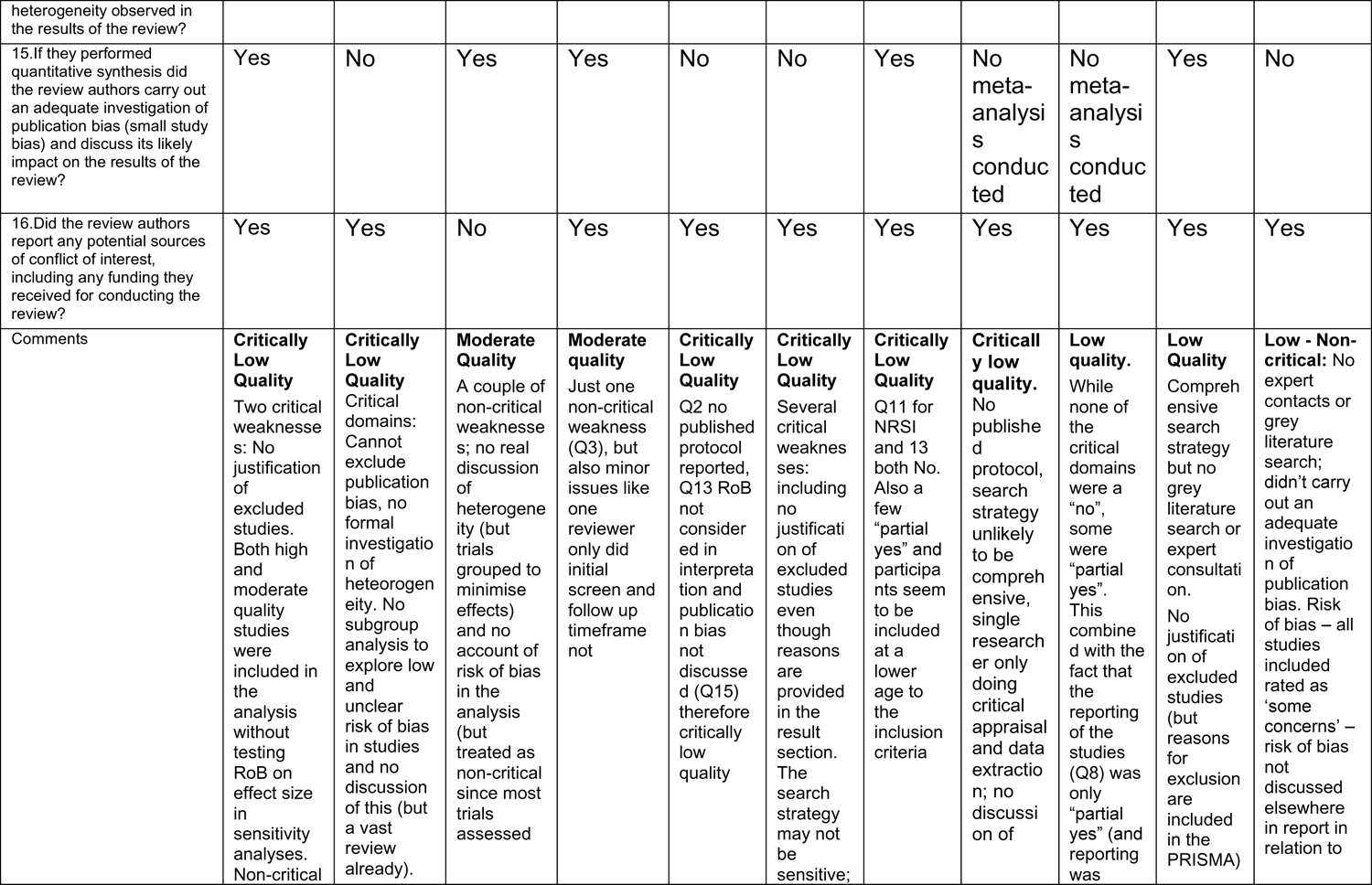
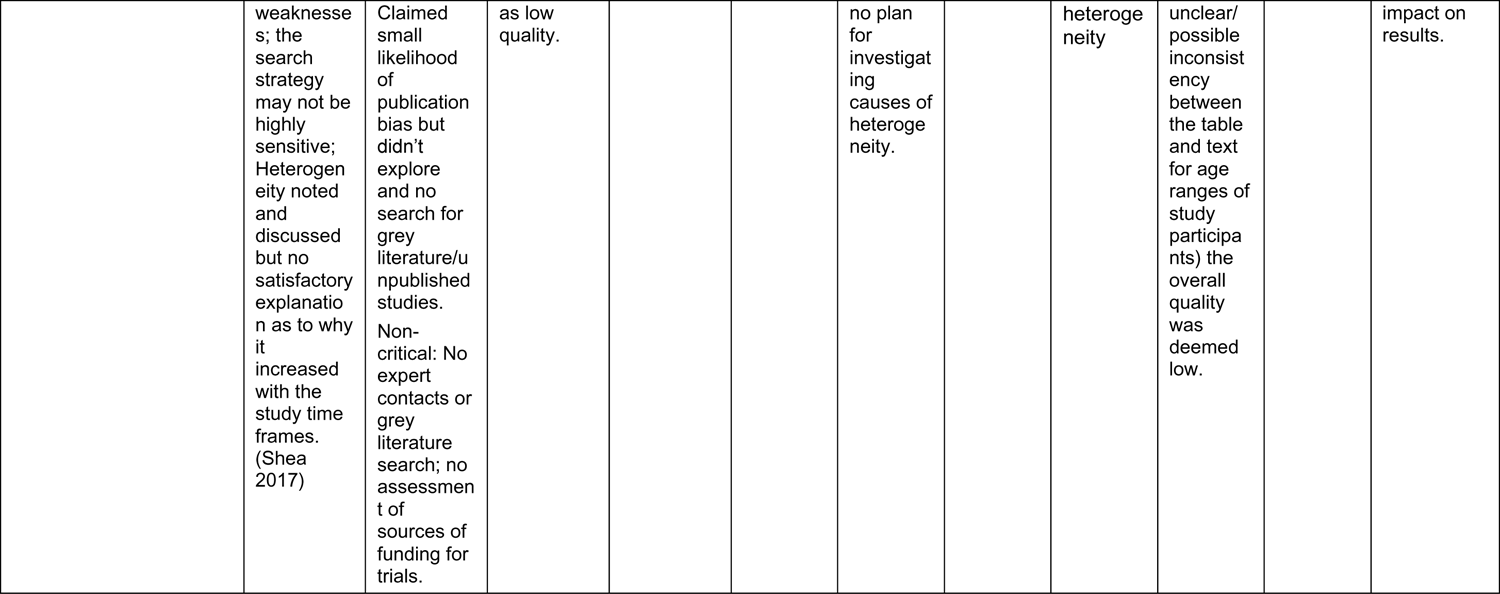

6.3 Quality appraisal of studies 6.3.1 Secondary research (Table 6)
| Item | Facchine<br>tti 2020 | Goncalve<br>s-Bradley<br>2017 | Goncalve<br>s-Bradley<br>2022 | Handoll<br>2021 | Langho<br>rne<br>2017 | Lee<br>2022 | Mabire<br>2016 | Meulen<br>broeks<br>2021 | Rassm<br>ussen | Tomlins<br>on 2020 | Williams<br>2022 |
| --- | --- | --- | --- | --- | --- | --- | --- | --- | --- | --- | --- |
| 1. Did the research questions and inclusion criteria for the review include the components of PICO? | Yes | Yes | Yes | Yes | Yes | Yes | Yes | Yes | Yes | Yes | Yes |
| 2. Did the report of the review contain an explicit statement that the review methods were established prior to the conduct of the review and did the report justify any significant deviations from the protocol? | Partial<br>Yes | Partial<br>Yes | Yes | Yes | No | Partial<br>Yes | Partial<br>Yes | No | Partial<br>Yes | Yes | Yes |
| 3. Did the review authors explain their selection of the study designs for inclusion in the review? | No | No | No | No | No | No | No | No | No | Yes | No |
| 4. Did the review authors use a comprehensive literature search strategy? | Partial<br>Yes | Partial<br>Yes | Yes | Yes | Yes | No | Partial<br>Yes | No | Partial<br>Yes | Partial<br>Yes | Partial<br>Yes |
| 5. Did the review authors perform study selection in duplicate? | Yes | Yes | Yes | Yes | Yes | No | Yes | No | Yes | Yes | Yes |
| 6. Did the review authors perform data extraction in duplicate? | Yes | Yes | Yes | Yes | Yes | Yes | Yes | No | Yes | Yes | Yes |
| 7. Did the review authors provide a list of excluded studies and justify the exclusion? | No | Yes | Yes | Yes | Yes | No | Partial<br>Yes | No | Yes | No | Yes |
| 8. Did the review authors describe the included studies in adequate detail? | Partial<br>Yes | Yes | Yes | Yes | Yes | Yes | Partial<br>Yes | Yes | Partial<br>Yes | Yes | Partial<br>Yes |
| 9. Did the review authors use a satisfactory technique for assessing the risk of bias (RoB) in individual studies that were included in the review? | Yes | Yes | Yes | Yes | Yes | Yes | RCTs;<br>Partial<br>Yes<br>NRSI;<br>Partial<br>Yes | RCTs;<br>Yes<br>NRSI;<br>Yes | RCTs;<br>Partial<br>Yes<br>NRSI;<br>Partial<br>Yes | Yes | RCTs;<br>Yes<br>NRSI; |
| 10. Did the review authors report on the sources of funding for the studies included in the review? | No | No | Yes | Yes | No | No | No | No | No | No | No |
| 11. If meta-analysis was performed did the review authors use appropriate methods for statistical combination of results? | RCTs;<br>Yes<br>NRSI;<br>No | No | RCTs;<br>Yes<br>NRSI;<br>No | RCTs;<br>Yes<br>NRSI;<br>No | RCTs;<br>Yes<br>NRSI;<br>No | No | Yes | No | No<br>meta-analysis<br>conduc<br>ted | RCTs;<br>Yes<br>NRSI; | RCTs;<br>Yes<br>NRSI; |
| 12. If meta-analysis was performed, did the review authors assess the potential impact of RoB in individual studies on the results of the meta-analysis or other evidence synthesis? | No | No | No | Yes | Yes | No | No | No<br>meta-analysis<br>conduc<br>ted | No<br>meta-analysis<br>conduc<br>ted | No | No |
| 13. Did the review authors account for RoB in individual studies when interpreting/discussing the results of the review? | No | Yes | No | Yes | No | No | No | Yes | Yes | Yes | No |
| 14. Did the review authors provide a satisfactory explanation for, and discussion of, any | No | No | No | Yes | Yes | No | Yes | No | Yes | Yes | Yes |
| heterogeneity observed in the results of the review? |  |  |  |  |  |  |  |  |  |  |  |
| 15.If they performed quantitative synthesis did the review authors carry out an adequate investigation of publication bias (small study bias) and discuss its likely impact on the results of the review? | Yes | No | Yes | Yes | No | No | Yes | No meta-analysis conducted | No meta-analysis conducted | Yes | No |
| 16.Did the review authors report any potential sources of conflict of interest, including any funding they received for conducting the review? | Yes | Yes | No | Yes | Yes | Yes | Yes | Yes | Yes | Yes | Yes |
| Comments | **Critically Low Quality**<br>Two critical weaknesses: No justification of excluded studies. Both high and moderate quality studies were included in the analysis without testing RoB on effect size in sensitivity analyses. Non-critical | **Critically Low Quality**<br>Critical domains: Cannot exclude publication bias, no formal investigation of heterogeneity. No subgroup analysis to explore low and unclear risk of bias in studies and no discussion of this (but a vast review already). | **Moderate Quality**<br>A couple of non-critical weaknesses; no real discussion of heterogeneity (but trials grouped to minimise effects) and no account of risk of bias in the analysis (but treated as non-critical since most trials assessed | **Moderate quality**<br>Just one non-critical weakness (Q3), but also minor issues like one reviewer only did initial screen and follow up timeframe not | **Critically Low Quality**<br>Q2 no published protocol reported, Q13 RoB not considered in interpretation and publication bias not discussed (Q15) therefore critically low quality | **Critically Low Quality**<br>Several critical weaknesses: including no justification of excluded studies even though reasons are provided in the result section. The search strategy may not be sensitive; | **Critically Low Quality**<br>Q11 for NRSI and 13 both No. Also a few "partial yes" and participants seem to be included at a lower age to the inclusion criteria | **Critically low quality.**<br>No published protocol, search strategy unlikely to be comprehensive, single researcher only doing critical appraisal and data extraction; no discussion of | **Low quality.**<br>While none of the critical domains were a "no", some were "partial yes". This combined with the fact that the reporting of the studies (Q8) was only "partial yes" (and reporting was | **Low Quality**<br>Comprehensive search strategy but no grey literature search or expert consultation. No justification of excluded studies (but reasons for exclusion are included in the PRISMA) | **Low - Non-critical:** No expert contacts or grey literature search; didn't carry out an adequate investigation of publication bias. Risk of bias – all studies included rated as 'some concerns' – risk of bias not discussed elsewhere in report in relation to |
|  | weaknesses; the search strategy may not be highly sensitive; Heterogeneity noted and discussed but no satisfactory explanation as to why it increased with the study time frames. (Shea 2017) | Claimed small likelihood of publication bias but didn't explore and no search for grey literature/unpublished studies. Non-critical: No expert contacts or grey literature search; no assessment of sources of funding for trials. | as low quality. |  |  | no plan for investigating causes of heterogeneity. |  | heterogeneity | unclear/possible inconsistency between the table and text for age ranges of study participants) the overall quality was deemed low. |  | impact on results. |

#### 6.3.2 Primary Research

##### Cohort studies (*Table 7*)

6.3.2 Primary Research *Cohort studies (Table 7)*
| Questions | Fisher 2020 |
| --- | --- |
| 1. Were the two groups similar and recruited from the same population? | Yes - from the United Kingdom Sentinel Stroke National Audit Programme |
| 2. Were the exposures measured similarly to assign people to both exposed and unexposed groups? | Yes - Early supported consensus score components |
| 3. Was the exposure measured in a valid and reliable way? | Yes – unique measure but based on 'international consensus and clinical guidelines' |
| 4. Were confounding factors identified? | Yes - rurality score and a hospital SSNAP rating score (total key indicator score derived across ten domains of stroke care with adjustments made for case ascertainment levels and the quality of data submitted to SSNAP) |
| 5. Were strategies to deal with confounding factors stated? | Yes – described on page 574 |
| 6. Were the groups/participants free of the outcome at the start of the study (or at the moment of exposure)? | Yes |
| 7. Were the outcomes measured in a valid and reliable way? | Yes - SSNAP audit |
| 8. Was the follow up time reported and sufficient to be long enough for outcomes to occur? | Yes, Jan 2016 to Dec 2016 |
| 9. Was follow up complete, and if not, were the reasons to loss to follow up described and explored? | Yes - explore the impact of missing data, we conducted sensitivity analysis excluding any teams that had missing outcome data; no substantial differences were found. |
| 10. Were strategies to address incomplete follow up utilized? | Yes |
| 11. Was appropriate statistical analysis used? | Yes |
| **Overall appraisal comments** | High quality, no areas of concern |

##### Cross-sectional studies (*Table 8*)

| Questions | Jones 2022 |
| --- | --- |
| 1. Were the criteria for inclusion in the sample clearly defined? | Yes – routinely collected data |
| 2. Were the study subjects and the setting described in detail? | Yes – routinely collected data |
| 3. Was the exposure measured in a valid and reliable way? | No – online questionnaire completed by discharge teams so open to bias |
| 4. Were objective, standard criteria used for measurement of the condition? | Yes – hospital data |
| 5. Were confounding factors identified? | Yes - after controlling for their characteristics, i.e. we identified DTOCs that could not be explained by LA-level demographic, demand and supply factors |
| 6. Were strategies to deal with confounding factors stated? | Yes - after controlling for their characteristics, i.e. we identified DTOCs that could not be explained by LA-level demographic, demand and supply factors |
| 7. Were the outcomes measured in a valid and reliable way? | Yes – RCD on DTOC |
| 8. Was appropriate statistical analysis used? | Yes |
| **Overall appraisal comments** | Moderate, potential bias in the way discharge teams completed questionnaire |

##### Qualitative studies (*Table 9*)

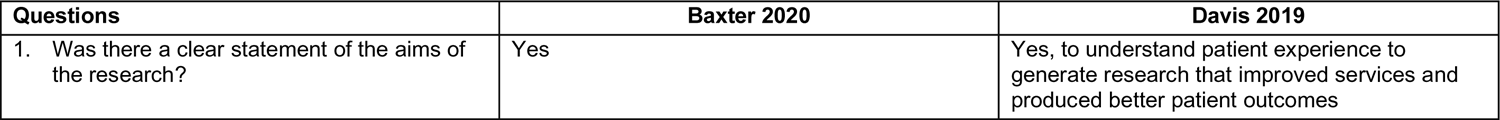
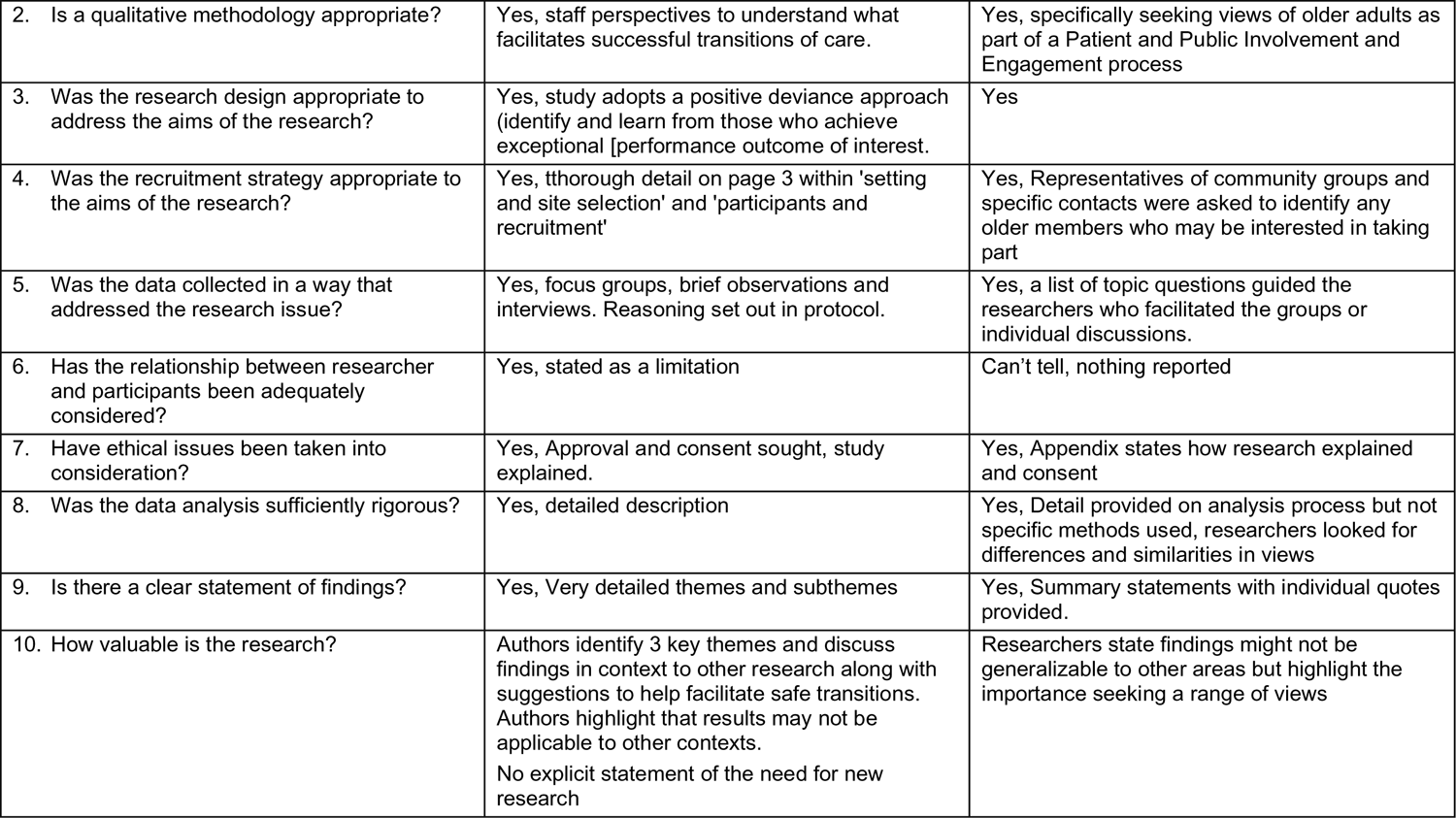

##### Quasi – experimental studies (*Table 10*)

| Questions | Elston 2022 |
| --- | --- |
| 1. Is it clear in the study what is the 'cause' and what is the 'effect' (i.e. there is no confusion about which variable comes first)? | Yes – investigating enhanced intermediate care (EIC) |
| 2. Were the participants included in any comparisons similar? | Unclear – limited details of population demographics of comparison areas |
| 3. Were the participants included in any comparisons receiving similar treatment/care, other than the exposure or intervention of interest? | Unclear – population level study so not clear other if other activities running at same time |
| 4. Was there a control group? | Yes |
| 5. Were there multiple measurements of the outcome both pre and post the intervention/exposure? | Yes- 2015, 2016 and 2017 but not clear when exactly data was collected in terms of introduction of EIC |
| 6. Was follow up complete and if not, were differences between groups in terms of their follow up adequately described and analyzed? | Not applicable – using service activity data |
| 7. Were the outcomes of participants included in any comparisons measured in the same way? | Yes |
| 8. Were outcomes measured in a reliable way? | Yes |
| 9. Was appropriate statistical analysis used? | Yes – not all data available to adjust for confounding such as 'package of care' |
| **Overall appraisal comments** | Moderate quality study, not all data is presented and lack of clarity in some areas of reporting. Not able to adjust for confounders and lack of demographic details of population. |

## 7. ADDITIONAL INFORMATION

### 7.1 Conflicts of interest

The authors declare they have no conflicts of interest to report.

## 7.2 Acknowledgements

The SURE team would like to thank stakeholders for their involvement in this rapid review process: Julie Rogers, Lisa Trigg, Sarah McCarty and Angie Oliver.

## 7.3 Disclaimer

The views expressed in this publication are those of the authors, not necessarily Health and Care Research Wales. The WCEC and authors of this work declare that they have no conflict of interest.

## 8. ABOUT THE WALES COVID-19 EVIDENCE CENTRE (WCEC)

The WCEC integrates with worldwide efforts to synthesise and mobilise knowledge from research.

We operate with a core team as part of Health and Care Research Wales, are hosted in the [Wales Centre for Primary and Emergency Care Research (PRIME)](http://www.primecentre.wales/), and are led by [Professor Adrian Edwards of Cardiff University](https://www.cardiff.ac.uk/people/view/123022-edwards-adrian).

The core team of the centre works closely with collaborating partners in [Health Technology Wales](https://www.healthtechnology.wales/), Wales Centre for Evidence-Based Care, [Specialist Unit for Review Evidence centre](https://www.cardiff.ac.uk/specialist-unit-for-review-evidence), [SAIL Databank](https://healthandcareresearchwales.org/about-research-community/secure-anonymised-information-linkage-sail-databank), Bangor Institute for Health & Medical Research/ Health and Care Economics Cymru, and the [Public Health Wales Observatory](https://phw.nhs.wales/services-and-teams/observatory/).

Together we aim to provide around 50 reviews per year, answering the priority questions for policy and practice in Wales as we meet the demands of the pandemic and its impacts.

## Director

Professor Adrian Edwards

## Website

https://healthandcareresearchwales.org/about-research-community/wales-covid-19-evidence-centre

## 9. APPENDICES

### Appendix 1: Resources searched during Rapid Review Searching

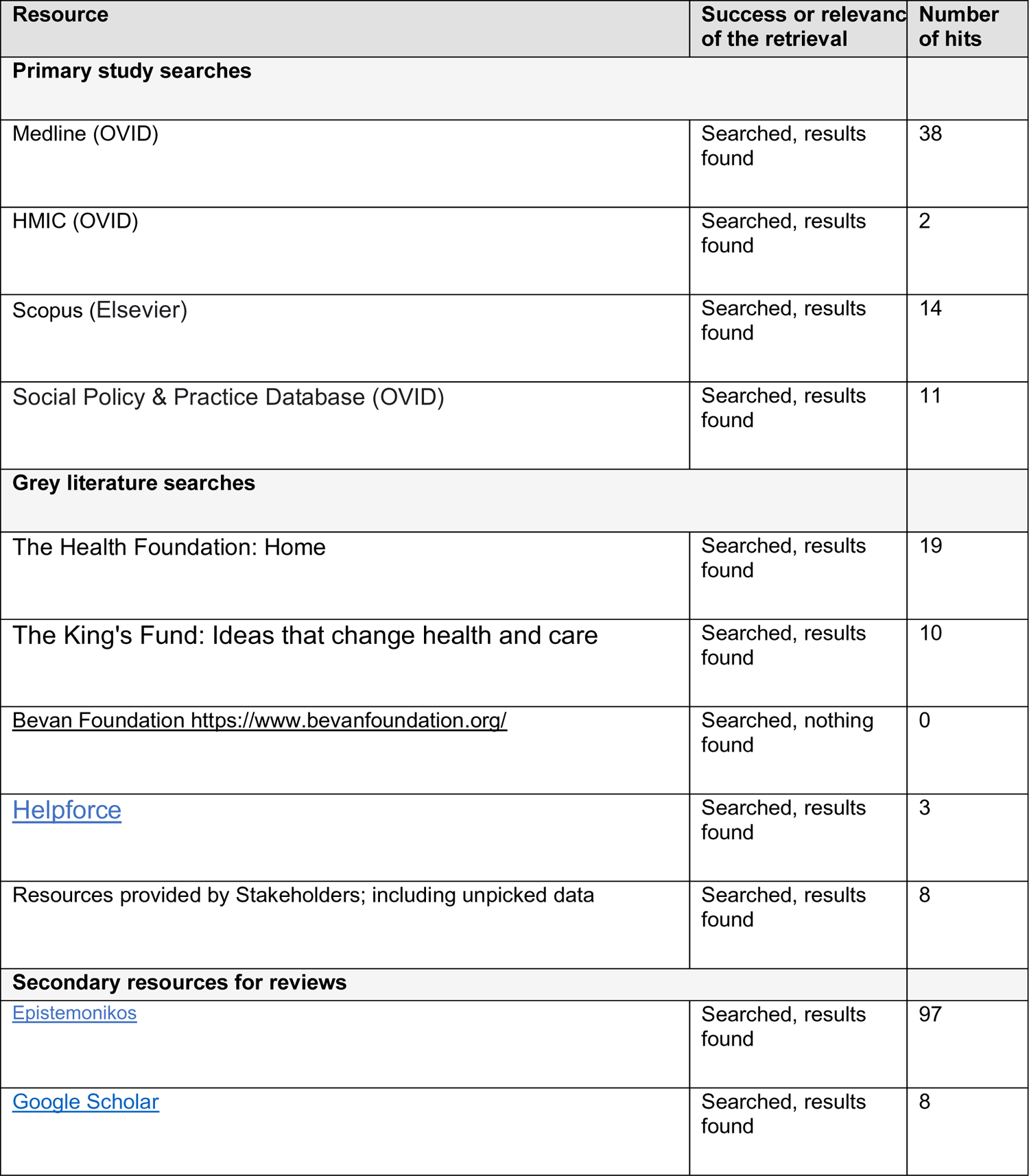

## Appendix 2: Search strategies

### Epistemonikos scoping search strategy

(title:((discharg* OR leav*) AND (hospital) AND (older OR elderly)) OR abstract:((discharg* OR leav*) AND (hospital) AND (older OR elderly))) Limit publication type: **systematic review** and 2020-2022.

### Medline search strategy

Ovid MEDLINE(R) ALL <1946 to May 20, 2022>

1. “Aged, 80 and over”/ 1006284
2. Aged/ 3350310
3. Frail Elderly/ 13901
4. (Late life or elder* or aged or old age or geriatric or seniors).tw. 946855
5. ((old or older or aging or aged or senior or elder*) adj3 (person or persons or people or adult* or patient* or male or males or female* or men or women)).tw. 924103
6. or/1-5 4218688
7. ((“medically fit” or “clinically optimi?ed”) adj3 discharge).tw. 23
8. ((time* or early or earlier or prompt or accelerate* or acute or subacute or supported or assisted) adj3 discharg*).tw. 20359
9. ((early or earlier or prompt or accelerate* or supported) adj4 return* adj2 home*).tw. 61
10. (leave adj3 hospital).tw. 960
11. 7 or 8 or 9 or 10 21349
12. (early supported discharge or ESD).tw. 5355
13. (discharge to assess or discharge to recover).tw. 173
14. exp Home Care Services/ 49977
15. “hospital rehabilitation unit*”.tw. 53
16. ((post-discharge or home rehabilitation) adj5 (support* or care)).tw. 966
17. (hospital* adj3 home*).tw. 12303
18. (rehabilitation adj3 home*).tw. 2788
19. (intensive adj2 home adj5 (rehabilitation or support*)).tw. 31
20. “organi?ed home care”.tw. 21
21. (mobile adj2 team*).tw. 763
22. ((community or nursing home or care home or domiciliary or primary care or home or home-based) adj3 (rehabilitation or support* or care)).tw. 193655
23. ((Intermediate or transition*) adj2 care).tw. 8271
24. (supportive care or rehabilitative care).tw. 18717
25. ((home first or safely home or step down) adj3 care).tw. 194
26. integrated care.tw. 5374
27. overseas recruitment.tw. 40
28. (redeploy* adj5 staff).tw. 129
29. (realign* adj5 team*).tw. 6
30. ((staff or team* or workforce) adj5 (upskilling or reskilling)).tw. 56
31. (multidisciplinary team* or MDT).tw. 24490
32. workforce model*.tw. 156
33. patient reablement.tw. 0
34. or/12-33 291097
35. exp United Kingdom/ 384740
36. (national health service* or nhs*).ti,ab,in. 244612
37. (english not ((published or publication* or translat* or written or language* or speak* or literature or citation*) adj5 english)).ti,ab. 44552
38. (gb or “g.b.” or britain* or (british* not “british columbia”) or uk or “u.k.” or united kingdom* or (england* not “new england”) or northern ireland* or northern irish* or scotland* or scottish* or ((wales or “south wales”) not “new south wales”) or welsh*).ti,ab,jw,in. 2308140
39. (bath or “bath’s” or ((birmingham not alabama*) or (“birmingham’s” not alabama*) or bradford or “bradford’s” or brighton or “brighton’s” or bristol or “bristol’s” or carlisle* or “carlisle’s” or (cambridge not (massachusetts* or boston* or harvard*)) or (“cambridge’s” not (massachusetts* or boston* or harvard*)) or (canterbury not zealand*) or (“canterbury’s” not zealand*) or chelmsford or “chelmsford’s” or chester or “chester’s” or chichester or “chichester’s” or coventry or “coventry’s” or derby or “derby’s” or (durham not (carolina* or nc)) or (“durham’s” not (carolina* or nc)) or ely or “ely’s” or exeter or “exeter’s” or gloucester or “gloucester’s” or hereford or “hereford’s” or hull or “hull’s” or lancaster or “lancaster’s” or leeds* or leicester or “leicester’s” or (lincoln not nebraska*) or (“lincoln’s” not nebraska*) or (liverpool not (new south wales* or nsw)) or (“liverpool’s” not (new south wales* or nsw)) or ((london not (ontario* or ont or toronto*)) or (“london’s” not (ontario* or ont or toronto*)) or manchester or “manchester’s” or (newcastle not (new south wales* or nsw)) or (“newcastle’s” not (new south wales* or nsw)) or norwich or “norwich’s” or nottingham or “nottingham’s” or oxford or “oxford’s” or peterborough or “peterborough’s” or plymouth or “plymouth’s” or portsmouth or “portsmouth’s” or preston or “preston’s” or ripon or “ripon’s” or salford or “salford’s” or salisbury or “salisbury’s” or sheffield or “sheffield’s” or southampton or “southampton’s” or st albans or stoke or “stoke’s” or sunderland or “sunderland’s” or truro or “truro’s” or wakefield or “wakefield’s” or wells or westminster or “westminster’s” or winchester or “winchester’s” or wolverhampton or “wolverhampton’s” or (worcester not (massachusetts* or boston* or harvard*)) or (“worcester’s” not (massachusetts* or boston* or harvard*)) or (york not (“new york*” or ny or ontario* or ont or toronto*)) or (“york’s” not (“new york*” or ny or ontario* or ont or toronto*))))).ti,ab,in. 1620778
40. (bangor or “bangor’s” or cardiff or “cardiff’s” or newport or “newport’s” or st asaph or “st asaph’s” or st davids or swansea or “swansea’s“).ti,ab,in. 64752
41. (aberdeen or “aberdeen’s” or dundee or “dundee’s” or edinburgh or “edinburgh’s” or glasgow or “glasgow’s” or inverness or (perth not australia*) or (“perth’s” not australia*) or stirling or “stirling’s“).ti,ab,in. 238977
42. (armagh or “armagh’s” or belfast or “belfast’s” or lisburn or “lisburn’s” or londonderry or “londonderry’s” or derry or “derry’s” or newry or “newry’s“).ti,ab,in. 30955
43. or/35-42 2897716
44. (exp africa/ or exp americas/ or exp antarctic regions/ or exp arctic regions/ or exp asia/ or exp australia/ or exp oceania/) not (exp United Kingdom/ or europe/) 3200959
45. 43 not 44 2746264
46. 6 and 11 and 34 and 45 235
47. limit 46 to (english language and yr=“2019 - 2022“) 38

## Footnotes

1 Lefebvre C, Glanville J, Briscoe S, Littlewood A, Marshall C, Metzendorf M-I, Noel-Storr A, Rader T, Shokraneh F, Thomas J, Wieland LS. Chapter 4: Searching for and selecting studies. In: Higgins JPT, Thomas J, Chandler J, Cumpston M, Li T, Page MJ, Welch VA (editors). Cochrane Handbook for Systematic Reviews of Interventions version 6.2 (updated February 2021). Cochrane, 2021. Available at: https://training.cochrane.org/handbook/current/chapter-04 [Accessed: 08 June 2022]

2 Based on available definitions, the following interventions were group under Early Supported Discharge: Early Discharge Hospital at Home/Discharge to Assess/Home Based Bridging/Home First

3 Welsh Government. Home First: The Discharge to Recover then Assess model (Wales). A summary guide to the principles and process. Cardiff: Welsh Government, December 2021 https://gov.wales/sites/default/files/publications/2022-01/the-discharge-to-recover-then-assess-model.pdf

4 Department of Health & Social Care. Hospital Discharge and Community Support Guidance. London: DHSC, March 2022 Hospital Discharge and Community Support Guidance (publishing.service.gov.uk)

## Notes

### Competing Interest Statement

The authors have declared no competing interest.

### Funding Statement

The Specialist Unit for Review Evidence was funded for this work by the Wales Covid-19 Evidence Centre, itself funded by Health & Care Research Wales on behalf of Welsh Government.

